# Not all movements are equal: Differences in variability of trunk motor behavior between people with and without low back pain - A Systematic Review

**DOI:** 10.1101/2023.06.06.23290554

**Authors:** Florian Abu Bakar, J. Bart Staal, Robert van Cingel, Hiroki Saito, Raymond Ostelo, Jaap H. van Dieën

## Abstract

**Background:** In treatment of low-back pain (LBP), motor control exercises have shown to be superior to minimal interventions, but not to any other form of exercise therapy. Knowledge about variability in trunk motor behavior may help to identify patients that may be more likely to benefit from motor control exercises.

**Objective:** This systematic review aims to answer the question: Is variability of trunk motor behavior different between people with and without LBP and if so, do people with LBP show more or less variability? Furthermore, we addressed the question whether the results are dependent on characteristics of the patient group, the task performed and the type of variability measure.

**Methods:** This study was registered in PROSPERO (CRD42020180003). Studies were eligible if they (1) included a LBP group and a control group, (2) included adults with non-specific low back pain of any duration and (3) measured kinematic variability, EMG variability and/or kinetic variability. Risk of Bias was evaluated and a descriptive synthesis was performed.

**Results:** Thirty-nine studies were included, thirty-one of which were included in the descriptive synthesis. In most studies and experimental conditions, variability did not significantly differ between groups. When significant differences were found, less variability in patients with LBP was more frequently reported than more variability, especially in gait-related tasks.

**Conclusions:** Given the considerable risk of bias of the included studies and the clinical characteristics of the participants with low severity scores for pain, disability and psychological measures, there is insufficient evidence to draw firm conclusions.

## Introduction

Low-back pain (LBP) is a widely prevalent condition causing a high burden of disease globally(1) and is associated with high economic costs(2). Unfortunately, effective treatment options are scarce(3). Regarding exercise therapy, many different treatment options in the management of chronic low-back pain (cLBP) have been studied. Exercise therapy is more effective for pain and disability than no treatment or usual care (4). However, there is limited evidence to support one type of exercise over another(3–6). Potential reasons for this are the heterogeneity of the patient group and a lack of knowledge regarding mechanisms that impede or facilitate recovery(7,8). Unraveling these mechanisms may facilitate improvement of cLBP-management(9).

Motor control exercises are commonly applied by physiotherapists in the treatment of LBP(10). Compared to minimal intervention (i.e., placebo intervention, education or advice and no treatment), motor control exercises have shown to be superior, but as other treatment modalities with modest effect sizes. Moreover, motor control exercises do not seem to be superior to any other form of exercise therapy(10–12). This might suggest that some (subgroups of) patients do benefit from motor control exercise while others do not or to a lesser degree. The identification of LBP patients with motor control alterations who are more likely to benefit from motor control exercises, as well as the appropriate choice of an individualized exercise regimen remains challenging due to lack of evidence (13,14).

Human motor behavior is characterized by substantial variation and variability in motor output. In this context, ‘variation’ refers to the array of movement possibilities a person has in everyday life to achieve a movement goal (e.g., walking to the store versus running versus going by bike) while ‘variability’ refers to differences within the same movement (e.g., within the walking pattern when walking to the store)(15,16). In other words, variability refers to the variance that occurs across multiple repetitions of the same movement (e.g., between strides taken while walking), which are never repeated in exactly the same manner(17–19) or to variance that occurs in static postures. Whether this variance is due to noise or determinism and even purposeful is debated(19,20). However, regardless the nature of the source of variability, its effect will be manifested in variance between motor outputs across repetitions of a movement at identical phases within this movement. When considering a static posture, this will be manifested in variance of motor outputs over time points.

There is an increasing body of literature suggesting differences in variability of trunk motor behavior between people with and without LBP (21,22). However, there seems to be no consistency in the direction of these alterations(13). Some studies reported less(23–25) variability in people with LBP while others reported more(26,27) variability or no difference at all(28). It could be hypothesized that different effects of cLBP on variability of trunk motor behavior could provide a basis for identifying patients who may be more likely to benefit from certain interventions such as motor control exercises. Zooming in on motor variability, motor outputs can be studied using kinematic measures (e.g., at the level of segment and joint movements), electromyography (e.g., at the level of muscle activation) or kinetic measures (e.g., at the level of muscle force production). Additionally, different methods to quantify variability in LBP have been applied. Some authors quantified the magnitude of variability in a set of measurements, expressed by measures such as standard deviation or range. Others advocated quantification of the structure of variability to assess the temporal organization in the distribution of the data, expressed by a large number of measures such as sample entropy (29) and the largest Lyapunov exponent (30). Yet, it is unclear if and how these different measures are related. In view of the lack of consensus in the literature regarding the operationalization of variability, this review will classify studies in 6 subgroups: Magnitude kinematic variability; Structure kinematic variability; Magnitude EMG variability; Structure EMG variability; Magnitude kinetic variability and Structure kinetic variability.

Without a comprehensive overview of the literature, it remains challenging to draw any conclusions regarding the association of LBP and variability in trunk motor behavior. Therefore, this systematic review aims to summarize and to synthesize current knowledge regarding this topic. More specifically, the objective is to answer the question: Is variability of trunk motor behavior different between people with and without LBP and if so, do people with LBP show more or less variability? Furthermore, the question is addressed whether the results are dependent on characteristics of the patient group, the task performed and the type of variability measure used.

## Methods

This systematic review was developed according to the Preferred Reporting Items of Systematic Reviews and Meta-Analyses Protocol (PRISMA-P) guideline(31,32) and has been registered in the PROSPERO database (CRD42020180003).

### Eligibility criteria

To be included in this review, the studies had to fulfill the following criteria:

#### Types of study

Studies that investigated both a LBP group and a healthy control group (cross-sectional as well as longitudinal) were included. Studies were excluded if they did not have a healthy control group, were a literature review of any kind or were animal studies. Articles in languages other than English were excluded.

#### Types of participants

Studies including adults (18 or older) with non-specific LBP, both acute (0-12 weeks) and chronic (>12 weeks) were included. Studies were excluded if participants had specific forms of LBP (e.g., fracture, infection, cancer, central nervous system disease, respiration disorders) or were post-surgery.

#### Types of outcome measures regarding variability

Studies were included if the construct to be measured was kinematic variability, EMG variability and/or kinetic variability. This was further subdivided into one of six subgroups of variability: Magnitude kinematic variability; Structure kinematic variability; Magnitude EMG variability; Structure EMG variability; Magnitude kinetic variability and Structure kinetic variability. Kinematic variability was defined as measurements of variability in kinematic outputs of the trunk (during movements/postures). EMG variability was defined as measurements of variability in trunk muscle activity as assessed with electromyography (during movement/postures). Kinetic variability was defined as measurements of variability in force exertion of the trunk, as assessed with inverse dynamics or dynamometry (during movement/postures).

### Search methods

A comprehensive systematic literature search was performed by the first author and an information specialist of our institution. Studies were identified by searching PubMed, Embase, Cinahl, Cochrane Central Register of Controlled Trials, Web of Science and Sport Discus from inception up till May 2022. The full search strategy for all databases can be seen in the supporting information. In addition, reference checking as well as citation checking of the included studies was performed to identify additional relevant studies.

### Study selection

The initial screening was performed by pairs of reviewers (FA-JvD, FA-RvC, FA-JBS, FA-RO) and consisted of applying the criteria for eligibility by screening the abstracts and titles retrieved by the search strategy(33). During all stages of the study selection process, disagreements were solved by discussion and consensus between the pairs of reviewers.

Where no consensus could be reached, a third reviewer of the group arbitrated. Where no abstract was available, full-text articles were obtained unless the article could be confidently excluded by its title alone. In general, if there was any doubt about the exclusion of a particular study, the study proceeded to full-text screening. For the application of the in-and exclusion criteria on the selected full text articles, the authors excluded an article when one of the exclusion criteria was met without registering the presence of additional exclusion criteria. Studies were classified as *included or excluded* using the web-tool Rayyan(34). As a group, the involved reviewers had relevant research experience in this field, were practicing clinicians or had extensive training in epidemiology, methodology or movement sciences.

### Data extraction

To decide on the content of the data to be extracted the checklist for critical appraisal and data extraction for systematic reviews of prediction modelling studies (CHARMS-PF) was used(35) as a guideline. The studies were classified into six subgroups. The data extraction from full texts was performed by one review author (FA). Two other authors (RO, JBS) verified the extraction table during the risk of bias assessment.

The following data were extracted:

*General study characteristics*: author, year of publication. *Clinical characteristics*: duration and severity of LBP. *Outcomes*: classes of tasks, variability measures and results.

### Risk of bias assessment

Risk of bias (RoB) was assessed with the Quality in Prognosis Studies (QUIPS) tool(36), a tool recommended by the Cochrane Methods Prognosis group(37). The QUIPS tool considers the following 6 domains of bias: Bias due to study participation, study attrition, prognostic factor measurement, outcome measurement, study confounding and statistical analysis & reporting.

Within each domain the three to seven items are usually scored. Possible responses were: ‘yes’, ‘partial’, ‘no’ or ‘unsure’. When specific information regarding items was not explicitly provided, we labeled it with ‘unsure’. The responses on these items were combined to assess the risk of bias per domain. The risk of bias for each domain was classified as ‘high’, ‘moderate’ or ‘low’(35). Regarding the domain ‘Study confounding’, it was decided that the highest achievable score was ‘moderate’ since comprehensive knowledge regarding confounders is lacking. ‘Moderate’ was scored when both, age and gender, were taken into account. Additionally, it was decided not to score the domain ‘Prognostic Factor Measurement’, since it did not differ from the domain ‘Study Participation’ in the context of this review. An overall risk of bias was not reported(38). Risk of bias was assessed by pairs of independent reviewers (RO, JBS, FA) (39). A priori, a calibration process was held for standardization purposes. After individual ratings, the results were compared. Disagreements were solved by discussion and consensus between reviewers.

### Analysis

Given the nature of the data, we refrained from doing a meta-analysis and performed a descriptive synthesis of the study results. Studies from the two kinematic subgroups (magnitude and structure) were included in this descriptive synthesis. Studies from the four other subgroups (i.e., magnitude and structure of EMG variability and kinetic variability) were not included due to heterogeneity in outcome measures and the low number of studies. Within the kinematic subgroups, four different classes of outcomes to measure variability were included, two for magnitude and two for structure. For magnitude measures, studies that measured variability in trunk angles (mean and standard deviation of trunk segments) or coordination (deviation phase or relative phase variability) were included. For structural measures, studies that used the short-term Lyapunov Exponent or %Determinism were included. Tasks were grouped into 5 classes of tasks; flexion-extension (consisting of trunk flexion-extension, lifting and sit-to-stand), gait (consisting of treadmill and overground walking and running), reaching, repositioning and static postures. Finally, the direction of the outcomes was considered. The possible outcomes were ‘no difference’, ‘more variability’ (i.e., magnitude: larger, structure: less regular in people with LBP) or ‘less variability’ (i.e., magnitude: smaller, structure: more regular in people with LBP). In the latter two cases (i.e., ‘more variability’ or ‘less variability’) between group differences should be statistically significant.

In the descriptive synthesis, experimental conditions from the two kinematic subgroups were taken into account and presented in tables. Each table row shows the experimental condition with outcomes (i.e., ‘less variability’, ‘no difference’ and ‘more variability’) within the five groups of tasks. The description of the distribution of all experimental conditions identified in the literature gives an indication of the direction of the reported results. For example, when the overall distribution of the results is more towards less variability, one might tentatively conclude that variability is reduced on average in the patients, since null findings in individual studies or experimental conditions do not provide evidence for absence of a difference between groups. On the other hand, when the outcomes are symmetrically distributed, with similar numbers of studies showing more and less variability, this strongly suggests that on average the groups do not differ. Additionally, the study references per cell are shown.

## Results

### Literature search

A total of 3802 articles were identified in the search after duplicates had been removed. This included six articles that were included after additional reference and citation checking. These articles were screened for eligibility based on title and abstract. This resulted in the exclusion of 3569 articles. The remaining 233 articles were screened for eligibility based on the full text. Finally, a total of 39 articles were included in this systematic review (*Fig 1*).

**Fig 1.**
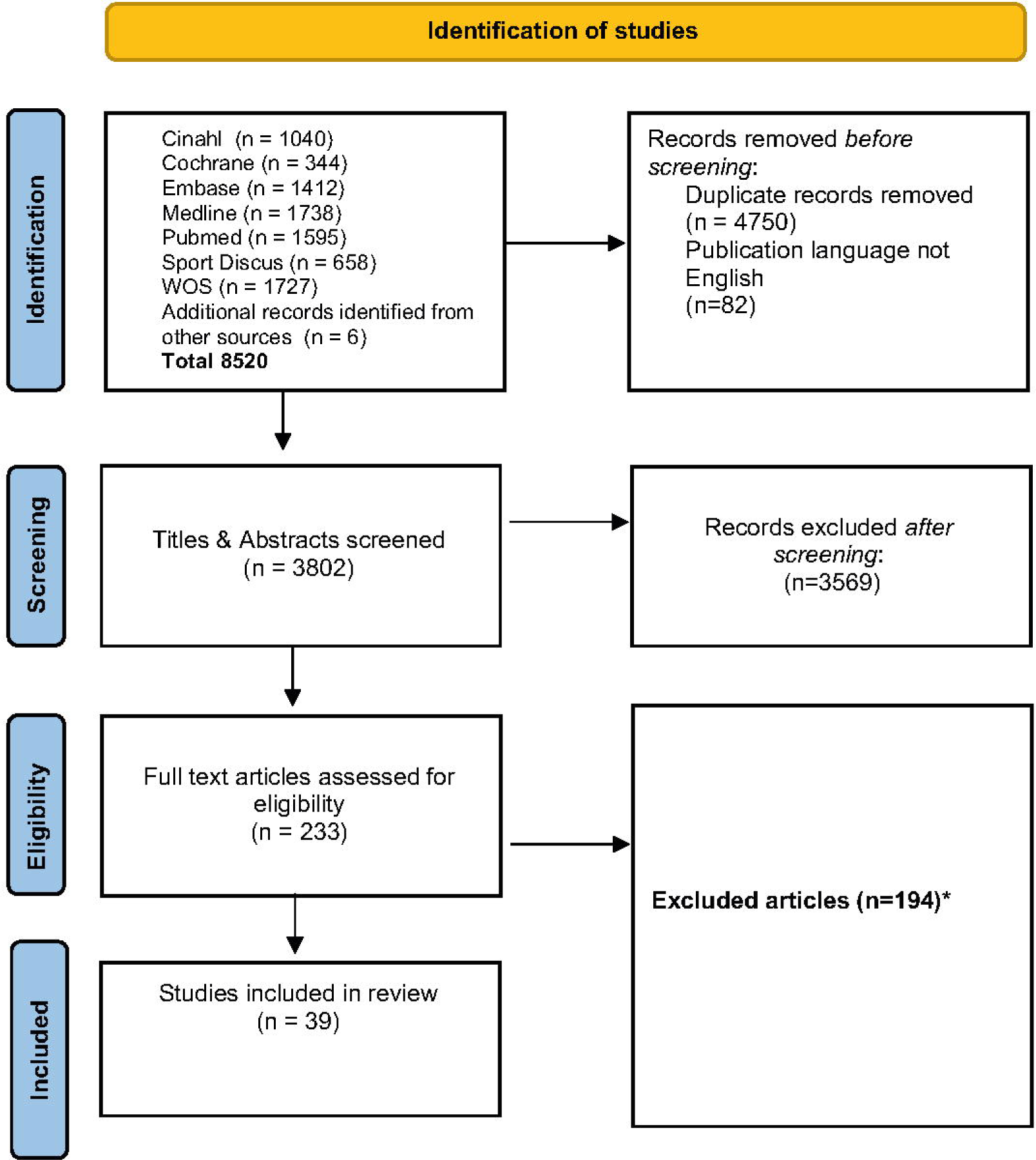
Flowchart of study inclusion in the systematic review. *Full text articles were excluded when one of the exclusion criteria was met without registering the presence of additional exclusion criteria.

### Study characteristics

The extracted data can be seen in tables 1-5. Thirty-nine studies, with 1486 participants (754 with LBP and 732 controls) were included in this review. All but one study(40) had a cross-sectional study design. Sample sizes for patient groups varied from 4 to 63 with an average of 19 subjects. Sample sizes for control groups varied from 6 to 80 with an average of 19 subjects. Twenty-one of the thirty-nine studies (22,40–59) matched participants with and without LBP on the following factors: sex (n=21), age (n=14), body mass/Body Mass Index (n=8), height (n=5) and level of activity (n=3).

**Table 1.** Studies regarding Magnitude of kinematic variability.

| Author | Year | Clinical characteristics: Duration LBP in month | Clinical characteristics: Pain | Clinical characteristics: Disability | Clinical characteristics: Psychological | Classes of tasks | Variability Measure | Direction: More variability-Less variability-No Difference |
| --- | --- | --- | --- | --- | --- | --- | --- | --- |
| Asgari, M | 2015 | *not reported* | *not reported* | *not reported* | *not reported* | Flexion-Extension | Mean&SD of coefficient of variation and variance ratio | No difference |
| Bagheri, R | 2020 | *not reported* | NPRS 5,5 | ODI 12,4 | *not reported* | Gait | Mean&SD of pattern variability and offset variability | Less (pattern)<br>No difference (offset) |
| Descarreaux, M | 2005 | 48,9 | VAS 15,5 | ODI 27,4 | *not reported* | (M)VC of trunk muscles | Mean&SD of peak angular position variability | No difference |
| Dideriksen, J | 2014 | 34,2 | NPRS 3,1 | ODI 14,2 | TSK 31,8<br>PCS 16,1<br>SF-36 66,9<br>STAI 40,2 | Flexion-Extension | Mean&SD of spinal angles/angular trajectories | No difference |
| Dunk, N | 2010 | *not reported* | *not reported* | ODI 15,9 | *not reported* | Static Postures | Mean&SD of lumbar movement patterns | More |
| Ebrahimi, S | 2017 | *not reported* | NPRS 5,1 | ODI 37,3 | *not reported* | Gait | Mean&SD of DP | Less |
| Fujii, R | 2022 | 14,7 | NPRS 3,6 | RDQ 2,1 | TSK-short 21,9<br>PCS-short 6,4 | Flexion-Extension | Mean&SD of DP | No difference |
| Hamacher, D | 2014 | *not reported* | *not reported* | *not reported* | *not reported* | Gait | Mean&SD of stride-to-stride variability | More (dual task)<br>No difference (single task) |
| Ippersiel, P | 2018 | 109,9 | NPRS 3,4 | ODI 25,5 | SBT 4,4 | Flexion-Extension | Mean&SD of DP | More |
| Lamoth, C (A) | 2006 | *not reported* | *not reported* | *not reported* | *not reported* | Gait | Mean & SD of relative phase, residual pattern variability | More (frontal plane)<br>Less (transv. plane) |
| Lamoth, C (B) | 2006 | *not reported* | VAS 53 | RDQ 10 | TSK 39 | Gait | Mean & SD of relative phase, residual pattern variability | More (frontal plane)<br>Less (transv. plane) |
| Mehravar, M | 2012 | 47 | VAS 10,9 | ODI 14,7 | *not reported* | Flexion-Extension | Mean&SEM of trunk segments | More |
| Mokhtarinia, H | 2016 | *not reported* | *not reported* | *not reported* | *not reported* | Flexion-Extension | Mean&SD of DP | No difference |
| Müller, J | 2017 | *not reported* | v Korff 50 | v Korff 41 | *not reported* | Gait | Mean&SD of coefficient of variation | No difference |
| Pelegrinelli, A | 2020 | 30 | VAS 6 | RDQ 2 | LoBACS 94 | Gait | Mean&SD of coupling angle variability | No difference |
| Ringheim, I | 2019 | *not reported* | NPRS 6 | ODI 26,9<br>PAL 7,4 | TSK 27,1 | Static Postures | Median&IQR of coefficient of variation | More |
| Rum, L | 2021 | *not reported* | VAS 46,5 | ODI 12,5 | TSK 30 | Gait | Mean & SD of AVgSD of angular displacements | More (transv. & frontal plane)<br>No difference (sagittal plane) |
| Seay, J | 2014 | *not reported* | VAS 8 | ODI 7,9 | *not reported* | Gait | Mean&SD of Relative Phase | No difference |
| Seay, J | 2011 | *not reported* | VAS 8 | ODI 7,9 | *not reported* | Gait | Mean&SD of Relative Phase | Less |
| Selles, R | 2001 | *not reported* | *not reported* | *not reported* | *not reported* | Gait | Mean&SD of Relative Phase | Less |
| Shojaei, I | 2020 | *not reported* | WBPI 5,2 | RDQ 11,8 | *not reported* | Flexion-Extension | Mean&SD of DP | Less |
| Shojaei, I | 2017 | *not reported* | *not reported* | *not reported* | *not reported* | Flexion-Extension | Mean&SD of DP | Less |
| Silfies, S | 2009 | 3,1 | NPRS 3,8 | RDQ 8,1 | *not reported* | Reaching | Mean&SD of DP | More |
| Tsigkanos, C | 2021 | *not reported* | *not reported* | *not reported* | *not reported* | Gait | Mean&SD lumbar angles | Less (No difference in most indices) |
| van den Hoorn, W | 2012 | 60 | VAS 29,2 | *not reported* | *not reported* | Gait | Mean&SD of stride-to-stride variability | Less |
| Wildenbeest, M | 2022 | *not reported* | NPRS 2,4 | ODI 31,4 | PCS 13<br>PASS 45<br>SBT 1,6<br>EBS 3 | Reaching | Mean&SD of lumbar angles and cycle times | No difference |

NPRS Numeric Pain Rating Scale, VAS Visual Analogue Scale, Korff Pain Intensity and Disability Scores , ODI Oswestry Disability Index, RDQ Roland Morris Disability Questionnaire, PAL Physical Activity Level, TSK Tampa Scale for Kinesiophobia, PCS Pain Catastrophizing Scale, SF-36 Short Form Health Survey, STAI State Trait Anxiety Inventory, SBT Start Back Tool, LoBACS Low Back Activity Confidence Scale, PASS Pain Anxiety Symptoms Scale, EBS Expected Back Strain Scale; DP Deviation Phase

**Table 2.**
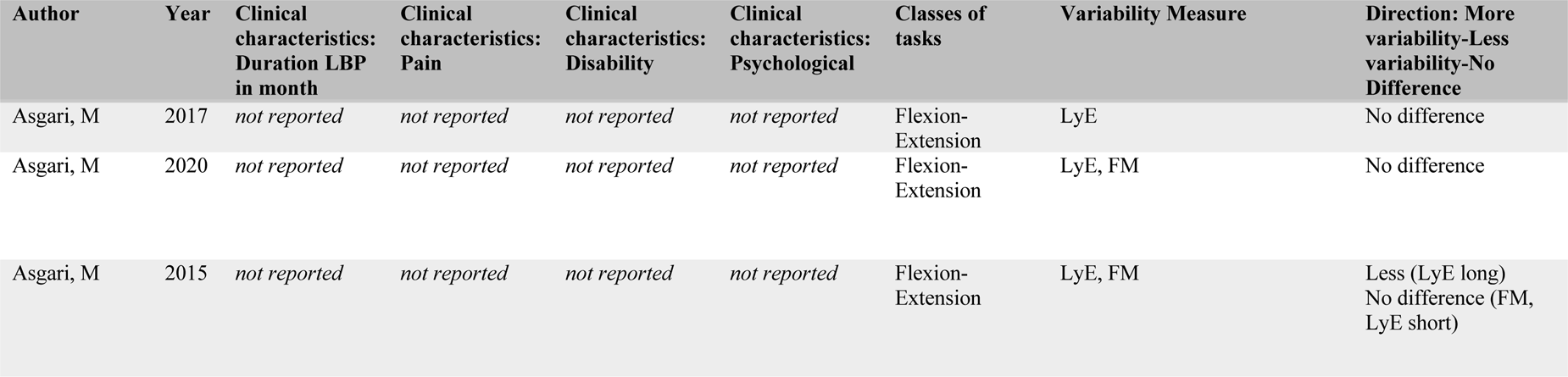
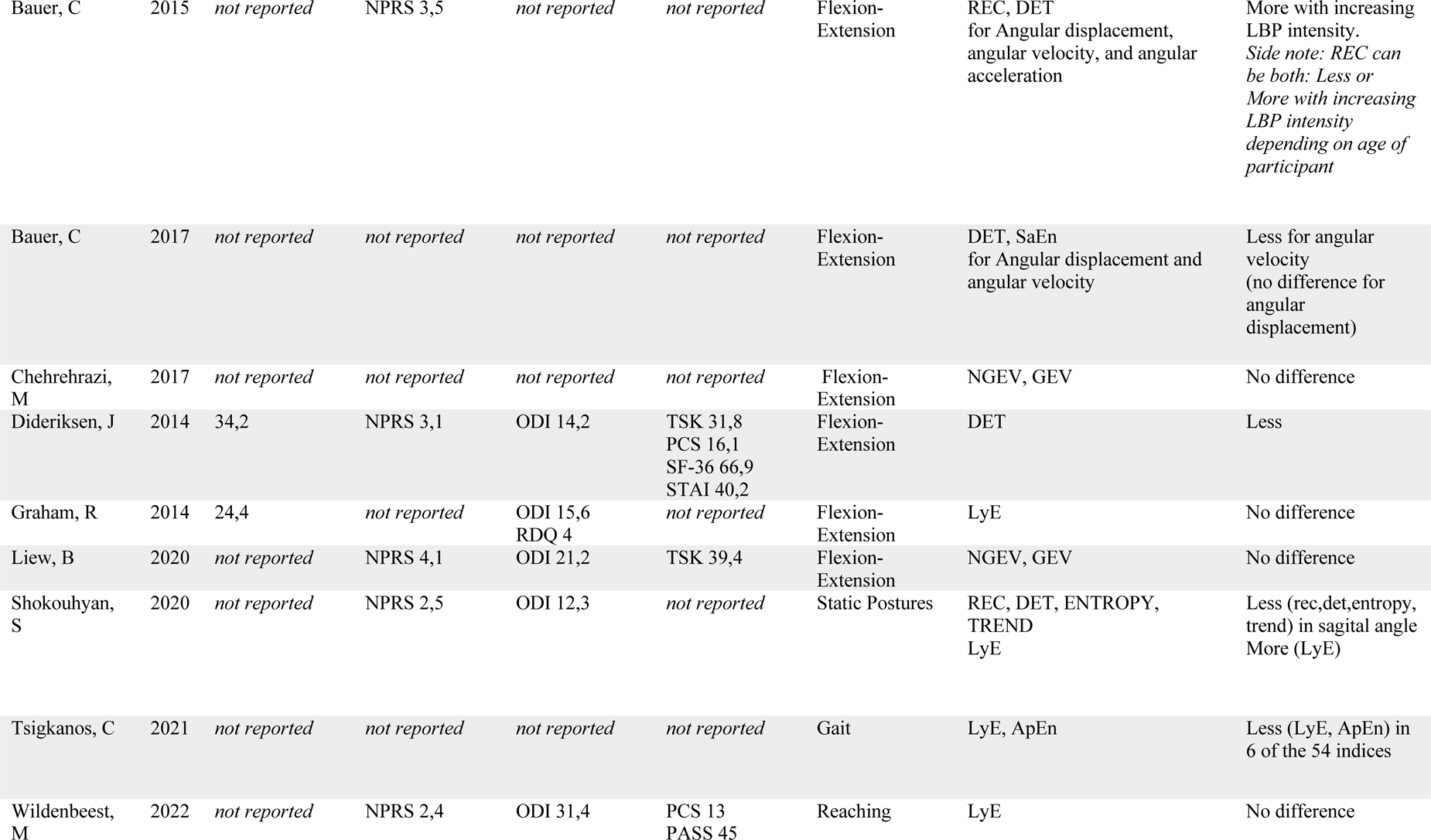
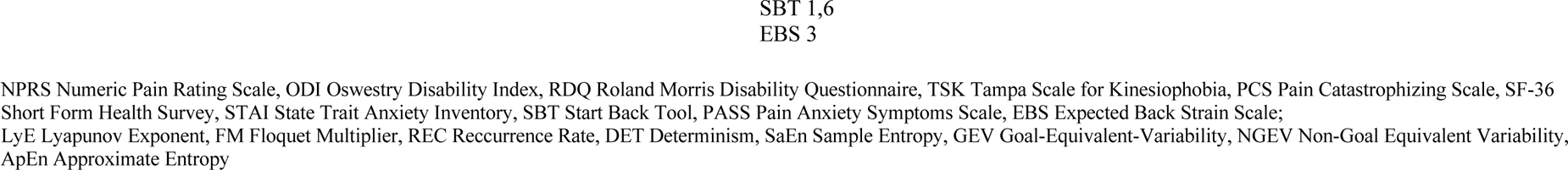
Studies regarding Structure of kinematic variability.

**Table 3.** Studies regarding Magnitude of EMG variability.

| Author | Year | Clinical characteristics: Duration LBP in month | Clinical characteristics: Pain | Clinical characteristics: Disability | Clinical characteristics: Psychological | Classes of tasks | Variability Measure | Direction: More variability-Less variability-No Difference |
| --- | --- | --- | --- | --- | --- | --- | --- | --- |
| Abboud, J | 2014 | *not reported* | VAS 46,5 | ODI 18,6 | TSK 36,3<br>SBT 2,8 | (M)VC of trunk muscles | Mean&SD of centre of gravity dispersion,<br>Changes of muscle recruitment pattern and intensity | Less |
| Arjunan, P | 2010 | *not reported* | VAS 35 | *not reported* | *not reported* | Gait | Change of variance of EMG amplitude over time | More |
| Falla, D | 2014 | 31,6 | NPRS 3,1 | ODI 13,8 | TSK 32,1<br>PCS 14,5<br>SF-36 67,6<br>STAI 40,1 | Flexion-Extension | RMS | Less |
| Jabobs, J | 2009 | *not reported* | NPRS 1,8 | ODI 13 | *not reported* | Static Postures | Variability of APA onset times | Less |
| Lamoth, C (A) | 2006 | *not reported* | *not reported* | *not reported* | *not reported* | Gait | Mean&SD of residual pattern variability | More |
| Lamoth, C (B) | 2006 | *not reported* | VAS 53 | RDQ 10 | TSK 39 | Gait | Mean&SD of residual pattern variability | More |
| Müller, J | 2017 | *not reported* | v Korff 50 | v Korff 41 | *not reported* | Gait | Coefficient of variation | No difference |
| Ringheim, I | 2019 | *not reported* | NPRS 6 | ODI 26,9<br>PAL 7,4 | TSK 27,1 | Static Postures | RMS | Less |

NPRS Numeric Pain Rating Scale, VAS Visual Analogue Scale, Korff Pain Intensity and Disability Scores , ODI Oswestry Disability Index, RDQ Roland Morris Disability Questionnaire, PAL Physical Activity Level, TSK Tampa Scale for Kinesiophobia, PCS Pain Catastrophizing Scale, SF-36 Short Form Health Survey, STAI State Trait Anxiety Inventory, SBT Start Back Tool; RMS Root Mean Square

**Table 4.** Studies regarding Structure of EMG variability.

| Author | Year | Clinical characteristics: Duration LBP in month | Clinical characteristics: Pain | Clinical characteristics: Disability | Clinical characteristics: Psychological | Classes of tasks | Variability Measure | Direction: More variability-Less variability-No Difference |
| --- | --- | --- | --- | --- | --- | --- | --- | --- |
| Graham, R | 2014 | 24,4 | *not reported* | ODI 15,6<br>RDQ 4 | *not reported* | Flexion-Extension | LyE | More |
| Liew, B | 2020 | *not reported* | NPRS 4,1 | ODI 21,2 | TSK 39,4 | Flexion-Extension | NGEV, GEV | No difference |

NPRS Numeric Pain Rating Scale, VAS Visual Analogue Scale, ODI Oswestry Disability Index, RDQ Roland Morris Disability Questionnaire, TSK Tampa Scale for Kinesiophobia, LyE Lyapunov Exponent, GEV Goal-Equivalent-Variability, NGEV Non-Goal Equivalent Variability

**Table 5.** Studies regarding Magnitude of kinetic variability.

| Author | Year | Clinical characteristics: Duration LBP in month | Clinical characteristics: Pain | Clinical characteristics: Disability | Clinical characteristics: Psychological | Classes of tasks | Variability Measure | Direction: More variability-Less variability-No Difference |
| --- | --- | --- | --- | --- | --- | --- | --- | --- |
| Descarreaux, M | 2007 | *not reported* | VAS 20 | ODI 22,6 | *not reported* | Repositioning | Peak force variability | No difference |

VAS Visual Analogue Scale, ODI Oswestry Disability Index

### Clinical characteristics

There is a lack of information regarding the exact duration of LBP. Twenty-nine of the 39 studies (74%) did not report the duration. In other words, for 578 out of the 754 (77%) LBP participants this information was not available. For the remaining ten studies with 176 participants with LBP (23% of total) the average LBP duration was 40.4 months (SD 29.6). Regarding gender distribution, 566 (38%) of the 1486 LBP participants were female and 782 (53%) were male. Due to non-reporting, the gender distribution remains unclear in 138 (9%) LBP participants divided over four studies(40,60–62). The mean baseline pain level measured with the Visual Analog Scale (VAS, 0-100) was 25.3 (SD17.5). This was measured in 175 of the 754 LBP participants (23.2%), divided over 11 studies(42,49,56,62–69).

The mean baseline pain level measured with the Numeric Rating Scale (NRS, 0-10) was 3.7 (SD1.2). This was measured in 295 of the 754 LBP participants (39.1%), divided over 13 studies(22,40,46,47,51,52,54,59,70–74). The mean disability level measured with the Oswestry Disability Index (ODI, 0-100) was 18.5 (SD 8.1). This was measured in 328 of the 754 LBP participants (43.5%), divided over 19 studies(22,45,47,50–54,56,59,62–65,67,69– 72). The mean disability level measured with the Roland-Morris Questionnaire (RDQ, 0-24) was 6.3 (SD 4.2). This was measured in 123 of the 754 LBP participants (16.3%), divided over 6 studies(40,46,49,53,66,73). The mean kinesiophobia level measured with the Tampa scale (TSK, 17-68) was 34. This was measured in 141 of the 754 LBP participants (18.7%), divided over 7 studies(22,52,63,66,69,71,72). One study(46) used the short form of the TSK. The mean catastrophizing level measured with the Pain Catastrophizing Scale (PCS, 0-52) was 15. This was measured in 66 of the 754 LBP participants, divided over 3 studies(22,52,59). One study(46) used the short form of the PCS. Twenty-eight studies (27,40,42–45,47,48,50,51,53–58,60,62,64,65,67,68,73–78) representing 518 of the 754 LBP participants (68.7%) did not measure any psychological construct. Ten studies(27,43,44,48,55,60,75–78) representing 199 of the 754 LBP participants (26.4%) did not report the use of any questionnaires to characterize the sample.

The clinical characteristics of these participants point towards samples with relatively low levels of pain, low levels of disability and low scores in the varying psychological measures. Additionally, due to non-reporting, the clinical characteristics of 35% of the LBP participants remain unknown.

### Tasks and task characteristics

Eight type of trunk movement tasks were utilized to measure variability of trunk motor behavior. Treadmill walking and running tasks were most frequently used in ten studies(42,49,57,58,62,66–68,76,77), trunk flexion-extension in eight studies(40,43,44,48,53,55,78,79) and lifting in six studies (22,27,46,52,71,74). Other tasks were overground walking in four studies(47,51,60,69), static postures in four studies(45,50,54,72), reaching in two studies (59,73), sit-to-stand in two studies(56,70), (maximally) voluntary contractions of trunk muscles in two studies (63,65) and a repositioning task in one study(64).

### Outcome measures

Regarding the distribution of the included studies within the six subgroups, most studies (n=26) (22,40,43,46,47,49–51,55–60,62,64,66–70,72,73,76–78) focused on the *magnitude of kinematic variability*, followed by *structure of kinematic variability* (n=12) (22,27,43– 45,48,53,58,59,71,74,79) and *magnitude of EMG variability* (n=8) (42,52,54,57,63,66,72,76). Two studies(53,71) focused on the *structure of EMG variability*. One study(80) focused on the *magnitude of kinetic variability* and no study on the *structure of kinetic variability*.

### Risk of Bias

The Risk of Bias of all included studies is presented in *Table 6*. To reach consensus, 3 rounds were needed. Domains where first and second reviewers disagreed mainly concerned ‘Study Participation’, ‘Outcome Measurement’ and ‘Study Confounding’. Consultation of a third reviewer was necessary to resolve disagreement for 10.7% of all scores, mainly concerning domains ‘Outcome Measurement’ and ‘Statistical Analysis and Reporting’. There was no discernible difference in RoB within the subgroups or within the same reported direction of variability (i.e., less, more, no difference). In total, most items were scored as ‘moderate’ (53%), followed by ‘low’ (33%) and ‘high’ (13%). However, there were considerable differences in RoB within the 5 domains. Most of the studies had a low RoB in the domains ‘Outcome Measurement’ and ‘Statistical Analysis and Reporting’ (77% for both). The RoB of the domains ‘Study attrition’ and ‘Study Confounding’ was mostly rated moderate (90% and 92%). The RoB of domain ‘Study Participation’ was mostly rated moderate (51%) or high (46%).

**Table 6.** Risk of Bias of the included studies.

| Author | Year | Study Participation | Study Attrition | Prognostic Factor Measurement | Outcome Measurement | Study Confounding | Statistical Analysis and Reporting |
| --- | --- | --- | --- | --- | --- | --- | --- |
| Abboud, J | 2014 | Yellow | Yellow | Blue | Yellow | Yellow | Yellow |
| Arjunan, P | 2010 | Yellow | Yellow | Blue | Green | Yellow | Green |
| Asgari, M | 2015 | Yellow | Yellow | Blue | Green | Yellow | Green |
| Asgari, M | 2017 | Yellow | Yellow | Blue | Green | Yellow | Green |
| Asgari, M | 2020 | Yellow | Yellow | Blue | Green | Yellow | Green |
| Bagheri, R | 2020 | Yellow | Yellow | Blue | Green | Yellow | Green |
| Bauer, C | 2015 | Yellow | Yellow | Blue | Green | Yellow | Green |
| Bauer, C | 2017 | Yellow | Yellow | Blue | Green | Yellow | Green |
| Chehreghazi, M | 2017 | Red | Yellow | Blue | Green | Yellow | Green |
| Descarreux, M | 2005 | Red | Yellow | Blue | Red | Yellow | Yellow |
| Descarreaux, M | 2007 | High risk of bias | Medium risk of bias | not scored | High risk of bias | Medium risk of bias | Medium risk of bias |
| Dideriksen, J | 2014 | Medium risk of bias | Medium risk of bias | not scored | Low risk of bias | Medium risk of bias | Low risk of bias |
| Dunk, N | 2010 | High risk of bias | Medium risk of bias | not scored | Low risk of bias | Medium risk of bias | Low risk of bias |
| Ebrahimi, S | 2017 | Medium risk of bias | Medium risk of bias | not scored | Low risk of bias | Medium risk of bias | Low risk of bias |
| Falla, D | 2014 | Medium risk of bias | Medium risk of bias | not scored | Low risk of bias | Medium risk of bias | Low risk of bias |
| Fujii, R | 2022 | Low risk of bias | Medium risk of bias | not scored | Low risk of bias | Medium risk of bias | Low risk of bias |
| Graham, R | 2014 | Medium risk of bias | Medium risk of bias | not scored | Low risk of bias | Medium risk of bias | Low risk of bias |
| Hamacher, D | 2014 | High risk of bias | Medium risk of bias | not scored | Low risk of bias | High risk of bias | High risk of bias |
| Ippersiel, P | 2018 | Medium risk of bias | Medium risk of bias | not scored | Low risk of bias | Medium risk of bias | Medium risk of bias |
| Jabobs, J | 2009 | High risk of bias | Low risk of bias | not scored | Low risk of bias | Medium risk of bias | Low risk of bias |
| Lamoth, C (A) | 2006 | Medium risk of bias | Low risk of bias | not scored | Low risk of bias | Medium risk of bias | Low risk of bias |
| Lamoth, C (B) | 2006 | Medium risk of bias | Low risk of bias | not scored | Low risk of bias | Medium risk of bias | Low risk of bias |
| Liew, B | 2020 | High risk of bias | Low risk of bias | not scored | Low risk of bias | Medium risk of bias | Low risk of bias |
| Mehravar, M | 2012 | Medium risk of bias | Medium risk of bias | not scored | Medium risk of bias | Medium risk of bias | Low risk of bias |
| Mokhtarinia, H | 2016 | High risk of bias | Medium risk of bias | not scored | Low risk of bias | Medium risk of bias | Low risk of bias |
| Müller, J | 2017 | Medium risk of bias | Medium risk of bias | not scored | Medium risk of bias | Medium risk of bias | Low risk of bias |
| Pelegrielli, A | 2020 | High risk of bias | Medium risk of bias | not scored | Medium risk of bias | Medium risk of bias | Low risk of bias |
| Ringheim, I | 2019 | Medium risk of bias | Medium risk of bias | not scored | Low risk of bias | Medium risk of bias | Low risk of bias |
| Rum, L | 2021 | High risk of bias | Medium risk of bias | not scored | Low risk of bias | Medium risk of bias | Low risk of bias |
| Seay, J | 2011 | High risk of bias | Medium risk of bias | not scored | Low risk of bias | High risk of bias | High risk of bias |
| Seay, J | 2014 | High risk of bias | Medium risk of bias | not scored | Low risk of bias | High risk of bias | High risk of bias |
| Selles, R | 2001 | High risk of bias | Medium risk of bias | not scored | Low risk of bias | Medium risk of bias | Low risk of bias |
| Shojaei, I | 2017 | High risk of bias | Medium risk of bias | not scored | High risk of bias | Medium risk of bias | Low risk of bias |
| Shojaei, I | 2020 | High risk of bias | Medium risk of bias | not scored | High risk of bias | Medium risk of bias | Low risk of bias |
| Shokouhyan, S | 2020 | High risk of bias | Medium risk of bias | not scored | Low risk of bias | Medium risk of bias | Medium risk of bias |
| Silfies, S | 2009 | High risk of bias | Medium risk of bias | not scored | High risk of bias | Medium risk of bias | Low risk of bias |
| Tsigkanos, C | 2021 | High risk of bias | Medium risk of bias | not scored | Low risk of bias | Medium risk of bias | Medium risk of bias |
| van den Hoorn, W | 2012 | Medium risk of bias | Medium risk of bias | not scored | Low risk of bias | Medium risk of bias | Low risk of bias |
| Wildenbeest, M | 2022 | Medium risk of bias | Medium risk of bias | not scored | Low risk of bias | Medium risk of bias | Low risk of bias |

*Medium risk of bias* *Low risk of bias* *High risk of bias* *not scored*

### Analysis

Seventy-seven % of all participants were included in the descriptive synthesis, representing 31 of the 39 studies. Eight studies(42,48,52,54,63,65,71,74) were excluded because synthesis was not feasible due to variance in outcome measures or the low number of studies. These studies measured EMG variability and kinetic variability. This synthesis was based on 8247 observations (n experimental conditions x n subjects) and comprised 20 comparisons for 5 experimental tasks and 4 outcome measures. Variability of trunk motor behavior was not consistently different between people with and without LBP. For six comparisons, no data were available. In the remaining 14 comparisons, the most frequent observation was no between-group differences (10 times), five times with an even distribution between more and less variability. Two comparisons consistently indicated less variability in the LBP group and one comparison indicated more variability in the LBP group. A-symmetric distributions of outcomes suggesting less variability occurred 5 times and a-symmetric distributions indicating more variability occurred 3 times (tables 7 – 10).

**Table 7.**
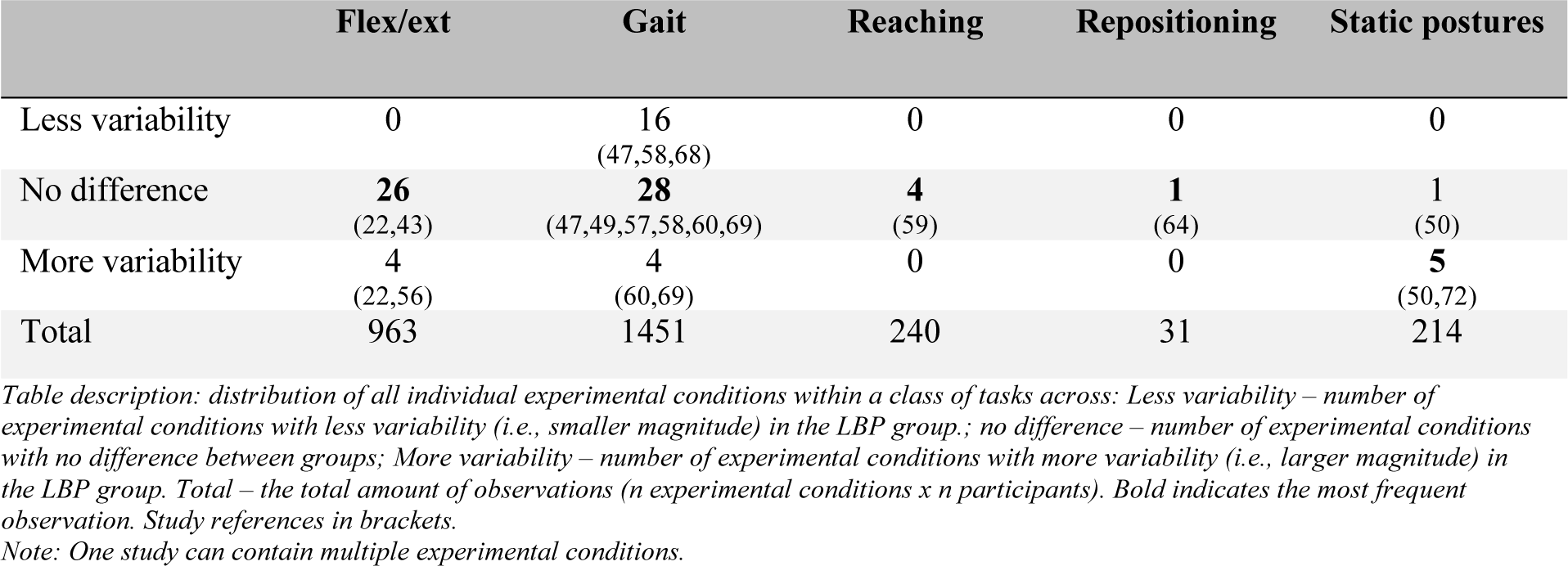
Magnitude trunk angles.

|  | Flex/ext | Gait | Reaching | Repositioning | Static postures |
| --- | --- | --- | --- | --- | --- |
| Less variability | 0 | 16<br>(47,58,68) | 0 | 0 | 0 |
| No difference | **26**<br>(22,43) | **28**<br>(47,49,57,58,60,69) | **4**<br>(59) | **1**<br>(64) | 1<br>(50) |
| More variability | 4<br>(22,56) | 4<br>(60,69) | 0 | 0 | **5**<br>(50,72) |
| Total | 963 | 1451 | 240 | 31 | 214 |

Table description: distribution of all individual experimental conditions within a class of tasks across: Less variability – number of experimental conditions with less variability (i.e., smaller magnitude) in the LBP group.; no difference – number of experimental conditions with no difference between groups; More variability – number of experimental conditions with more variability (i.e., larger magnitude) in the LBP group. Total – the total amount of observations (n experimental conditions x n participants). Bold indicates the most frequent observation. Study references in brackets. Note: One study can contain multiple experimental conditions.

**Table 8.** Magnitude coordination.

|  | Flex/ext | Gait | Reaching | Repositioning | Static postures |
| --- | --- | --- | --- | --- | --- |
| Less variability | **33**<br>(40,55,66,76,78) | 13<br>(51,67,77) | 0 | 0 | 0 |
| No difference | 23<br>(40,46,55,76,81) | **21**<br>(62,67) | 2<br>(82) | 0 | 0 |
| More variability | 5<br>(76,81) | 0 | 2<br>(82) | 0 | 0 |
| Total | 2341 | 824 | 260 | 0 | 0 |

Table description: distribution of all individual experimental conditions within a class of tasks across: Less variability – number of experimental conditions with less variability (i.e., smaller magnitude) in the LBP group.; no difference – number of experimental conditions with no difference between groups; More variability – number of experimental conditions with more variability (i.e., larger magnitude) in the LBP group. Total – the total amount of observations (n experimental conditions x n participants). Bold indicates the most frequent observation. Study references in brackets. Note: One study can contain multiple experimental conditions.

**Table 9.** Structure Lyapunov.

|  | Flex/ext | Gait | Reaching | Repositioning | Static postures |
| --- | --- | --- | --- | --- | --- |
| Less variability | 0 | 1<br>(58) | 0 | 0 | **2**<br>(45) |
| No difference | **14**<br>(27,43,44,53) | **8**<br>(58) | **1**<br>(59) | 0 | 0 |
| More variability | 0 | 0 | 0 | 0 | 0 |
| Total | 454 | 261 | 60 | 0 | 80 |

Table description: distribution of all individual experimental conditions within a class of tasks across: Less variability – number of experimental conditions with less variability (i.e., more regular structure) in the LBP group.; no difference – number of experimental conditions with no difference between groups; More variability – number of experimental conditions with more variability (i.e., less regular structure) in the LBP group. Total – the total amount of observations (n experimental conditions x n participants). Bold indicates the most frequent observation. Study references in brackets.
Note: One study can contain multiple experimental conditions.

**Table 10.**
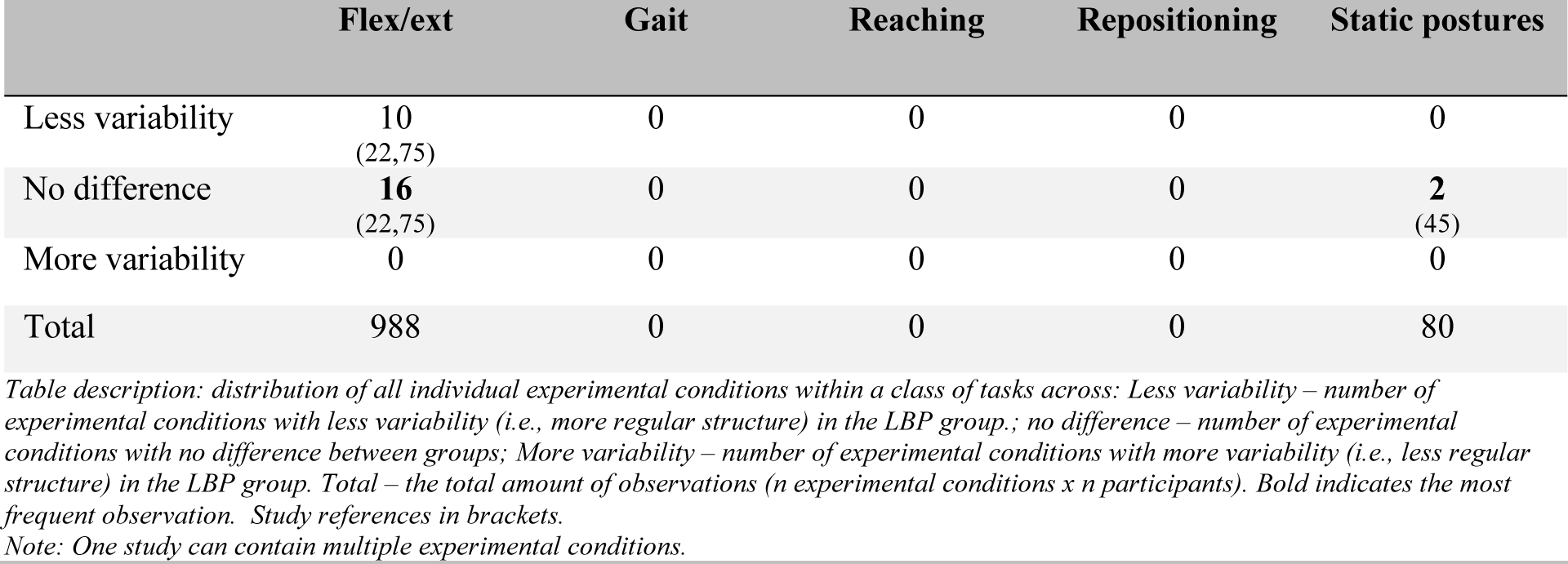
Structure Determinism.

|  | Flex/ext | Gait | Reaching | Repositioning | Static postures |
| --- | --- | --- | --- | --- | --- |
| Less variability | 10<br>(22,75) | 0 | 0 | 0 | 0 |
| No difference | **16**<br>(22,75) | 0 | 0 | 0 | **2**<br>(45) |
| More variability | 0 | 0 | 0 | 0 | 0 |
| Total | 988 | 0 | 0 | 0 | 80 |

Table description: distribution of all individual experimental conditions within a class of tasks across: Less variability – number of experimental conditions with less variability (i.e., more regular structure) in the LBP group.; no difference – number of experimental conditions with no difference between groups; More variability – number of experimental conditions with more variability (i.e., less regular structure) in the LBP group. Total – the total amount of observations (n experimental conditions x n participants). Bold indicates the most frequent observation. Study references in brackets.
Note: One study can contain multiple experimental conditions.

The secondary research questions focused on whether results were dependent on characteristics of the patient group, the tasks performed and the types of measurement that were used. Characteristics of the patient group could not be included in this descriptive synthesis because of low variance in clinical characteristics (e.g., pain, disability, and psychological measures), non-reporting of important characteristics in healthy controls, as well as too few data on general characteristics (e.g., age, duration of LBP).

Regarding the influence of the tasks, gait related tasks seemed to yield the most consistent differences, often showing no between-group difference, but when yielding a difference, it showed less variability of trunk motor behavior in patients with LBP and it did so across all measures where data were available (i.e., angles, coordination and Lyapunov exponent; no data for determinism). Flexion-extension tasks were less consistent, studies showed all possible outcomes (i.e., less variability, no differences and more variability) in participants with LBP. For example, regarding variability in magnitude of trunk angles (table 7) no between-group difference was reported most frequently, but when yielding a difference, it showed more variability of trunk motor behavior. Regarding variability in magnitude of coordination (table 8) less variability was reported most frequently, followed by no difference and more variability. Even though flexion-extension tasks were less consistent, no difference and less variability were more often found than more variability. For the other tasks (reaching, repositioning, static postures) it was not possible to draw conclusions due to the low number of observations.

Measures of magnitude seemed to be more sensitive than measures of structure. Trends in variability of trunk motor behavior towards more of less variability were more frequently reported for measures of magnitude (i.e., angles & coordination) than for measures of structure (i.e., Lyapunov & determinism), 6 out of 8 times and 3 out of 6 times respectively. The observations for magnitude measures were based on 26 studies and 937 participants, the observations for structure on 9 studies and 361 participants.

## Discussion

In this systematic review on variability of trunk motor behavior in low-back pain patients, studies were classified into one of six possible subgroups (i.e., magnitude kinematic variability; structure kinematic variability; magnitude EMG variability; structure EMG variability; magnitude kinetic variability and structure kinetic variability). Most studies focused on kinematic outcome measures. Studies from the 2 kinematic subgroups were used for a descriptive synthesis (table 7 – 10). The main research question of this review was whether variability differed between people with and without low back pain. We showed that, in most studies, variability did not differ between groups. Even though there was a substantial inconsistency regarding the direction of variability, less variability in patients with LBP was more frequently reported than more variability. Gait-related tasks often yielded no between-group difference, but when experimental tasks did yield a difference, it indicated less variability of trunk motor behavior in patients with LBP. The inconsistencies in variability of trunk motor behavior that was observed may be in agreement with the heterogeneous spectrum that LBP represents and has been reported before in reviews, in LBP(83,84) as well as in other populations(85–87).

In this review, trends towards less variability in patients with LBP was more frequently reported than towards more variability. This seems to be in line with van Dieën et al. 2017(88) who proposed a model in which changes in variability in LBP can be seen as a functional adaptation acquired through reinforcement learning. During the initial phase of LBP, variability might increase initially to decrease once a pattern has been found to control the pain and/or threat perceived. According to this suggestion, people with longstanding LBP will tend to control posture and movement more rigidly, causing more stereotypical muscle activation and kinematics. Even though there is a lack of reporting, most participants of this review seemed to have had chronic LBP with an average duration of 40 month.

The frequent occurrence of ‘no differences’ in variability between groups can have several reasons. There is a possibility that the pain, disability, and psychological measures in the group with LBP were not severe enough to detect significant differences between people with and without LBP. Potentially, the LBP participants of this review are not representative of a clinical LBP population. Most studies used open recruitment procedures (e.g., word of mouth, recruitment amongst university staff etc.) which might have led to the recruitment of people that experience LBP, but do not actively seek therapeutic guidance. Furthermore, larger between-subjects variance of trunk kinematics in populations with LBP compared to those without LBP(89) has been reported. This could lead to the representation of both groups (i.e., participants with more and participants with less variability) within one study. Finally, some studies had small sample size and/or small samples per subject to estimate within-subject variability in trunk motor behavior, which may have led to type 2 errors.

Concerning underlying mechanisms, both differences, more and less variability in LBP can be explained. More variability in LBP might be related to lower muscle activation or to proprioceptive impairments (13). Nociception has been shown to potentially cause reflexive inhibition of muscle activity (90), which would lead to lower muscle activity and consequently impaired control over joint movements. Nociception may also negatively affect proprioception (91), as has been confirmed for trunk proprioception and LBP (92). This would impair feedback control of trunk movement and as such could cause increased variability and decreased stability. On the other hand, less variability and an increase in stability might result from a protective movement strategy due to perceived or actual risk of pain provocation, potentially modulated by pain related cognitions or emotions (25). With acute pain, movement strategies are adapted to maintain control despite the disturbing effects of pain (e.g., the limping gait with small joint excursions after ankle injury). Such alterations in movement control involve high muscle activity and low variability in motor output (93). These divergent mechanisms might explain the inconsistency of the literature and indicate the necessity to diversify intervention approaches (94).

An additional question of this review was whether differences in trunk motor variability are dependent on the task performed. Upon visual inspection, the distribution of the individual experimental conditions within one outcome measure shows differences in direction of trunk variability between the different tasks. For example, regarding variability in magnitude of trunk angles (table 7), the overall distribution within gait-related tasks was towards less variability whereas within flexion-extension related tasks as well as within static postures it was towards more variability.

Besides differences in task, differences in task demands might have played a role as well. Unlike the other tasks, variability during gait-related tasks was generally measured at different speeds (i.e., walking and running), where higher speeds seemed to elicit a more consistent difference in variability, probably due to an increase of the task demand. Possible influences of task demands on variability have been reported before(84).

The influence of the patient characteristics could not be analyzed. Several characteristics (e.g., duration of complaints, description of pain, disability and psychological measures) remained largely unknown due to non-reporting (tables 1 – 5). Overall, the characteristics point towards samples with low levels of pain, low levels of disability and low scores (i.e., not deviating from normal) regarding various psychological measures. A possible explanation is that most studies were undertaken in movement labs and using open recruitment. Inclusion of participants with more severe and/or disabling forms of LBP in these kinds of settings might have been more challenging. A closer look at the few studies where LBP participants with higher (i.e., moderate) reported pain scores showed differences in variability in all cases, however, the direction of these differences still varied. Both directions, less variability (47,51,63,66,72) as well as more variability (66,69,72) were reported.

Regarding the risk of bias, several issues should be acknowledged. Most studies had a cross sectional design and from the longitudinal study(40) only baseline data were used. This implies that causal inferences cannot be made. The QUIPS domain of ‘Study participation’ was most frequently scored having a ‘high risk of bias’ (46% of all included studies). Regularly, essential information in this domain was lacking. Amongst others, this included information regarding the source population and the recruitment period and place. Furthermore, the domain ‘study attrition’ mostly scored a ‘moderate risk of bias’ (90%). On a regular basis, the information needed to score the items was not provided, resulting in an ‘unsure’ scoring of the item as agreed upon during the calibration process. Examples are missing information on the response rate and on the occurrence of loss to follow up. Concerning ‘study confounding’, most studies scored a ‘moderate risk of bias’. Due to the lack of knowledge regarding confounders for variability, it was decided that a low RoB score was not possible. By consensus, studies had to minimally report age and gender of the participants to score a moderate RoB. Most study findings represent merely non-adjusted, crude associations and generalization of the results should be taken cautiously. Maybe, this represents a certain lack of awareness of the construct of confounding in the included studies.

To ensure the methodological quality, the authors published a Prospero protocol (CRD42020180003) and followed Prisma guidelines(32). Nonetheless, there are limitations regarding the review processes. One limitation was that meta-analysis was deemed not feasible due to the nature of the data of the included studies. Therefore, statistical inference was not possible. However, a descriptive synthesis was employed to summarize and synthesize the data to answer the research questions. By doing so, 77% of the participants were represented in the descriptive synthesis. An expected finding was the variety in nomenclature and outcome measurements regarding variability(95). For example, nine of the thirty articles did not use the term ‘variability’ in title and abstract(27,40,44,45,50,53,74,77,78) and in the eight studies of the subgroup magnitude EMG(42,52,57,63,66,72,76) six different outcome measures were used to express variability. The authors anticipated this when constructing this review and conducted an extensive search of the literature based on a comprehensive search strategy developed by an information specialist with expertise in the field. This was supplemented with reference and citation checking. However, despite the systematic approach, the possibility exists that the used search terms did not comprise the large variety of measures and definitions that the construct of variability represents and therefore, relevant studies may have been missed. Variation between studies in movement tasks and movement task parameters (e.g., number of repetitions, standardization differences) was large. For example, of the sixteen studies within the subgroup of ‘flexion-extension’ twelve(22,40,46,52,56,64,65,71,74,75,78,81) employed a rather low number of repetitions to measure variability(96,97). This was considered during the RoB analysis. The use of the QUIPS tool in this review can be criticized, especially regarding the applicability and the arbitrary cut-off points. This was addressed by comparing concurrent tools with two experienced epidemiologists (JBS, RO) and by calibrating issues related to application and interpretation.

Recommendations for future studies include a clearer conceptualization as well as an operationalization of ‘variability’. While conceptual frameworks of variability have been described in the domains of sport and overuse injury(95,97), it remains challenging to transfer recommendations to the domain of LBP due to differences in scope. To achieve this, a broad consensus regarding definitions as well as parameters to employ when studying variability may be necessary. Future studies on variability should present rationales regarding issues like whether and which magnitude or structure measures are used, regarding the movement tasks (e.g., task complexity, the number of repetitions), as well as how to deal with possible confounders (e.g., age, gender, duration and severity of complaints, psychological factors, or endurance/fatigue related issues). Finally, it is recommended to include a broader spectrum of LBP participants (e.g., age, levels of pain and disability, psychological measures) and to provide complete description regarding the study participation and the clinical characteristics of the participants.

In conclusion, this systematic review provided an overview of the methods to measure variability in trunk motor behavior in people with LBP compared to people without and addressed the question whether variability of trunk motor behavior differed between them. In most of the studies and experimental conditions included, variability did not differ between groups, but when differences between groups were found, less variability in people with LBP was more frequently observed. Regarding implications for practice, as of now, a translation of the results towards the clinical setting is challenging and not recommended.

## Supporting information

S1 Checklist

S2 Checklist

S3 Checklist

## Data Availability

All relevant data are within the manuscript and its Supporting Information files.

## Acknowledgments

We thank Thomas Pelgrim (TP), information specialist at the HAN University of Applied Sciences, for his contribution to the search strategy.

## Authors Contributions

**Conceptualization**: Florian Abu Bakar, J.Bart Staal, Robert van Cingel, Raymond Ostelo, Jaap van Dieën.

**Data curation**: Florian Abu Bakar, J.Bart Staal.

**Formal analysis**: Florian Abu Bakar, Jaap van Dieën.

**Investigation**: Florian Abu Bakar.

**Methodology**: Florian Abu Bakar, J.Bart Staal, Robert van Cingel, Raymond Ostelo, Jaap van Dieën.

**Project administration**: Florian Abu Bakar.

**Supervision**: J.Bart Staal, Robert van Cingel, Raymond Ostelo, Jaap van Dieën.

**Writing – original draft**: Florian Abu Bakar.

**Writing – review & editing**: Florian Abu Bakar, J.Bart Staal, Robert van Cingel, Hiroki Saito, Raymond Ostelo, Jaap van Dieën.

## Supporting information

- **S1 Appendix. Search strategy for all databases.** (DOC)
- **S1 Checklist. Preferred reporting items for systematic review and meta-analysis (PRISMA) Abstract checklist.** (DOC)
- **S2 Checklist. Preferred reporting items for systematic review and meta-analysis (PRISMA) checklist.** (DOC)
- **S3 Checklist. Quality in Prognostic Studies (QUIPS) Tool.** (PDF)
- **S1 File. Registered protocol.** This is the review protocol which was registered with PROPERO. (PDF)

## Notes

### Competing Interest Statement

The authors have declared no competing interest.

### Funding Statement

Florian Abu Bakar was funded by a doctoral grand for teachers from the Dutch Organization for Scientific Research (NWO, grant number: 023.011.018). The other authors received no specific funding tor this work.

