## Supplementary material for "Not all movements are equal: Differences in variability of trunk motor behavior between people with and without low back pain - A Systematic Review": S1 Appendix

**Pubmed**

#1	“Low Back pain”[mesh] OR backpain*[tiab] OR lumbar pain*[tiab] OR lumbar back pain*[tiab] OR lumbar backach*[tiab] OR lumbar spine pain*[tiab] OR lbp[tiab] OR sacral pain*[tiab] OR dorsalgia[tiab] OR backach*[tiab] OR back ach*[tiab] OR back pain*[tiab] OR Lumbago*[tiab]

#2	“pain”[mesh] OR "Pain Measurement"[Mesh] OR pain[tiab] OR ache*[tiab] OR aching*[tiab] OR Physical Suffering*[tiab] OR Analges*[tiab] OR Nociception*[tiab]

#3	“Spine”[mesh] OR “back”[mesh] OR spine*[tiab] OR spinal[tiab]

#4	#2 AND #3

#5	#1 OR #4

#6	"Nonlinear Dynamics"[Mesh] OR Movement Variabilit*[tiab] OR Motor Variabilit*[tiab] OR Nonlinear Dynamic*[tiab] OR Non-linear Dynamic*[tiab] OR Nonlinear Model*[tiab] OR Non linear Models[tiab] OR Linear dynamic*[tiab] OR postural control[tiab] OR Postural sway[tiab] OR Trunk Control[tiab] OR Trunk muscle activ*[tiab] OR Movement irregularity[tiab] OR Movement alterations[tiab] OR Movement Variability[tiab] OR Coordinative Variability[tiab] OR Inter-segmental coordination [tiab] OR Trunk Pelvis coordination[tiab] OR Motor Variability[tiab] OR Kinetic Variability[tiab] OR Neuromuscular adaptations[tiab] OR Dynamical structure[tiab] OR Temporal structure[tiab] OR Temporal Spatial parameters[tiab] OR spine rotational stiffness[tiab] OR Dynamic stability[tiab] OR Trunk stability[tiab] OR trunk movement*[tiab] OR dynamic balance[tiab] OR kinematic variability*[tiab]

#7	"Entropy"[Mesh] OR Entrop*[tiab] OR Largest Lyapunov Exponent[tiab] OR detrended fluctuation analysis[tiab] OR correlation dimension[tiab] OR Mutual information[tiab] OR Hurst exponent[tiab] OR Recurrence quantification analysis[tiab] OR Floquet multiplier*[tiab]

#8	**"Electromyography"[Mesh] OR Center of Pressure[tiab] OR Vicon[tiab] OR OptiTrack[tiab] OR (**Optotrack[tiab] AND Certus[tiab]) OR Qualisys motion analysis system[tiab] OR (Liberty[tiab] AND Polhemus[tiab]) OR Electromagnetic motion tracking[tiab] OR Inertial Measurement Units[tiab] OR motion captur*[tiab]

**#9**	variabilit*[tiab] OR standard deviation[tiab] OR Range[tiab] OR excursion[tiab] OR Standard deviation[tiab] OR Mean[tiab] OR Angles[tiab] OR End point variability[tiab] OR Angular displacement[tiab] OR Acceleration[tiab] OR Velocity[tiab] OR Force accuracy[tiab] OR Total displacement[tiab]

**#10	#8 AND #9**

**#11	#6 OR #7 OR #10**

**#12	#5 AND #11**

**Medline**

S1	MH "Back Pain" OR MH "Low Back Pain" OR MH "Coccydynia" OR TI (backpain* OR ((lumbar OR back OR spine OR sacral OR spinal) N3 (pain* OR ache* OR aching OR (Physical N1 Suffering*) OR Analges* OR Nociception*)) OR lbp OR dorsalgia OR backach* OR Lumbago*) OR AB (backpain* OR ((lumbar OR back OR spine OR sacral OR spinal) N3 (pain* OR ache* OR aching OR (Physical N1 Suffering*) OR Analges* OR Nociception*)) OR lbp OR dorsalgia OR backach* OR Lumbago*)OR SU (backpain* OR ((lumbar OR back OR spine OR sacral OR spinal) N3 (pain* OR ache* OR aching OR (Physical N1 Suffering*) OR Analges* OR Nociception*)) OR lbp OR dorsalgia OR backach* OR Lumbago*)

S2	(MH "Pain+" OR MH "Pain Measurement") AND (MH "Spine+" OR MH "Back")

S3	S1 OR S2

S4	MH "Nonlinear Dynamics" OR TI (((Motor OR Movement OR kinetic OR kinematic) N1 Variabilit*) OR ((Nonlinear OR "Non linear" OR linear) N1 Dynamic*) OR ((Nonlinear OR "Non linear") N1 model) OR ((Trunk OR Postur*) N1 (control OR sway)) OR (Trunk N1 muscle* N1 activ*) OR (Movement N1 (irregularity OR alteration* OR Variabilit*)) OR (Coordinative N1 Variabilit*) OR ("Inter-segmental" N1 coordination) OR (Trunk N1 Pelvis N1 coordination*) OR (Neuromuscular N1 adaptation*) OR ((Dynamic* OR temporal) N1 structure*) OR (Temporal N1 Spatial N1 parameter*) OR (spine N1 rotational N1 stiffness) OR ((Trunk OR Dynamic) N1 (stability* OR movement OR balance))) OR AB (((Motor OR Movement OR kinetic OR kinematic) N1 Variabilit*) OR ((Nonlinear OR "Non linear" OR linear) N1 Dynamic*) OR ((Nonlinear OR "Non linear") N1 model) OR ((Trunk OR Postur*) N1 (control OR sway)) OR (Trunk N1 muscle* N1 activ*) OR (Movement N1 (irregularity OR alteration* OR Variabilit*)) OR (Coordinative N1 Variabilit*) OR ("Inter-segmental" N1 coordination) OR (Trunk N1 Pelvis N1 coordination*) OR (Neuromuscular N1 adaptation*) OR ((Dynamic* OR temporal) N1 structure*) OR (Temporal N1 Spatial N1 parameter*) OR (spine N1 rotational N1 stiffness) OR ((Trunk OR Dynamic) N1 (stability* OR movement OR balance))) OR SU (((Motor OR Movement OR kinetic OR kinematic) N1 Variabilit*) OR ((Nonlinear OR "Non linear" OR linear) N1 Dynamic*) OR ((Nonlinear OR "Non linear") N1 model) OR ((Trunk OR Postur*) N1 (control OR sway)) OR (Trunk N1 muscle* N1 activ*) OR (Movement N1 (irregularity OR alteration* OR Variabilit*)) OR (Coordinative N1 Variabilit*) OR ("Inter-segmental" N1 coordination) OR (Trunk N1 Pelvis N1 coordination*) OR (Neuromuscular N1 adaptation*) OR ((Dynamic* OR temporal) N1 structure*) OR (Temporal N1 Spatial N1 parameter*) OR (spine N1 rotational N1 stiffness) OR ((Trunk OR Dynamic) N1 (stability* OR movement OR balance)))

S5	MH "Entropy" OR TI (Entrop* OR (Largest N1 Lyapunov N1 Exponent*) OR (detrended N1 fluctuati* N1 analysis) OR (correlate* N1 dimension*) OR (Mutual N1 information) OR (Hurst N1 exponent*) OR (Recurrence N1 quantification N1 analysis) OR (Floquet N1 multiplier*)) OR AB (Entrop* OR (Largest N1 Lyapunov N1 Exponent*) OR (detrended N1 fluctuati* N1 analysis) OR (correlate* N1 dimension*) OR (Mutual N1 information) OR (Hurst N1 exponent*) OR (Recurrence N1 quantification N1 analysis) OR (Floquet N1 multiplier*)) OR SU (Entrop* OR (Largest N1 Lyapunov N1 Exponent*) OR (detrended N1 fluctuati* N1 analysis) OR (correlate* N1 dimension*) OR (Mutual N1 information) OR (Hurst N1 exponent*) OR (Recurrence N1 quantification N1 analysis) OR (Floquet N1 multiplier*))

S6	MH "Electromyography" **OR TI ((Center of Pressure*) OR Vicon OR OptiTrack OR (**Optotrack N1 Certus) OR (Qualisys N1 motion N1 analysis N1 system) OR (Liberty N1 Polhemus) OR (Electromagnetic N1 motion N1 track*) OR (Inertial N1 Measurement N1 Unit*) OR (motion N1 captur*)) **OR AB ((Center of Pressure*) OR Vicon OR OptiTrack OR (**Optotrack N1 Certus) OR (Qualisys N1 motion N1 analysis N1 system) OR (Liberty N1 Polhemus) OR (Electromagnetic N1 motion N1 track*) OR (Inertial N1 Measurement N1 Unit*) OR (motion N1 captur*)) **OR SU ((Center of Pressure*) OR Vicon OR OptiTrack OR (**Optotrack N1 Certus) OR (Qualisys N1 motion N1 analysis N1 system) OR (Liberty N1 Polhemus) OR (Electromagnetic N1 motion N1 track*) OR (Inertial N1 Measurement N1 Unit*) OR (motion N1 captur*))

**S7	TI (**variabilit* OR (standard N1 deviation*) OR Range OR excursion OR Mean OR Angles OR (End N1 point N1 variability*) OR (Angular N1 displac*) OR Acceleration* OR Velocit* OR (Force N1 accuracy*) OR (Total N1 displacement)) OR **AB (**variabilit* OR (standard N1 deviation*) OR Range OR excursion OR Mean OR Angles OR (End N1 point N1 variability*) OR (Angular N1 displac*) OR Acceleration* OR Velocit* OR (Force N1 accuracy*) OR (Total N1 displacement)) OR SU **(**variabilit* OR (standard N1 deviation*) OR Range OR excursion OR Mean OR Angles OR (End N1 point N1 variability*) OR (Angular N1 displac*) OR Acceleration* OR Velocit* OR (Force N1 accuracy*) OR (Total N1 displacement))

**S8	S6 AND S7**

**S9	S4 OR S5 OR S8**

**S10	S3 AND S9**

**Embase**

#1	'backache'/de or 'low back pain'/exp OR (backpain* OR ((lumbar OR back OR spine OR sacral OR spinal) ADJ3 (pain* OR ache* OR aching OR (Physical NEAR/1 Suffering*) OR Analges* OR Nociception*)) OR lbp OR dorsalgia OR backach* OR Lumbago*):ti,ab,kw

#2	'pain'/de or 'musculoskeletal pain'/de or 'spinal pain'/de OR 'pain measurement'/de

#3	'spine'/de or 'back'/de or 'lumbar spine'/de or 'lumbosacral spine'/de

#4	#2 AND #3

#5	#1 OR #4

#6	'nonlinear system'/de OR (((Motor OR Movement OR kinetic OR kinematic) NEAR/1 Variabilit*) OR ((Nonlinear OR "Non linear" OR linear) NEAR/1 Dynamic*) OR ((Nonlinear OR "Non linear") NEAR/1 model) OR ((Trunk OR Postur*) NEAR/1 (control OR sway)) OR (Trunk NEAR/1 muscle* NEAR/1 activ*) OR (Movement NEAR/1 (irregularity OR alteration* OR Variabilit*)) OR (Coordinative NEAR/1 Variabilit*) OR ("Inter-segmental" NEAR/1 coordination) OR (Trunk NEAR/1 Pelvis NEAR/1 coordination*) OR (Neuromuscular NEAR/1 adaptation*) OR ((Dynamic* OR temporal) NEAR/1 structure*) OR (Temporal NEAR/1 Spatial NEAR/1 parameter*) OR (spine NEAR/1 rotational NEAR/1 stiffness) OR ((Trunk OR Dynamic) NEAR/1 (stability* OR movement OR balance))):ti,ab,kw

#7	'entropy'/de OR (Entrop* OR (Largest NEAR/1 Lyapunov NEAR/1 Exponent*) OR (detrended NEAR/1 fluctuati* NEAR/1 analysis) OR (correlate* NEAR/1 dimension*) OR (Mutual NEAR/1 information) OR (Hurst NEAR/1 exponent*) OR (Recurrence NEAR/1 quantification NEAR/1 analysis) OR (Floquet NEAR/1 multiplier*)):ti,ab,kw

#8	'electromyography'/de OR **((Center of Pressure*) OR Vicon OR OptiTrack OR (**Optotrack NEAR/1 Certus) OR (Qualisys NEAR/1 motion NEAR/1 analysis NEAR/1 system) OR (Liberty NEAR/1 Polhemus) OR (Electromagnetic NEAR/1 motion NEAR/1 track*) OR (Inertial NEAR/1 Measurement NEAR/1 Unit*) OR (motion NEAR/1 captur*)):ti,ab,kw

**#9	(**variabilit* OR (standard NEAR/1 deviation*) OR Range OR excursion OR Mean OR Angles OR (End NEAR/1 point NEAR/1 variability*) OR (Angular NEAR/1 displac*) OR Acceleration* OR Velocit* OR (Force NEAR/1 accuracy*) OR (Total NEAR/1 displacement)):ti,ab,kw

**#10	#8 AND #9**

**#11	#6 OR #7 OR #10**

**#12	#5 AND #11**

**#13**	#12 AND 'conference abstract'/it

**#14	#12 NOT #13**

**Cinahl**

S1	MH "Back Pain" OR MH "Low Back Pain" OR MH "Coccydynia" OR TI (backpain* OR ((lumbar OR back OR spine OR sacral OR spinal) N3 (pain* OR ache* OR aching OR (Physical N1 Suffering*) OR Analges* OR Nociception*)) OR lbp OR dorsalgia OR backach* OR Lumbago*) OR AB (backpain* OR ((lumbar OR back OR spine OR sacral OR spinal) N3 (pain* OR ache* OR aching OR (Physical N1 Suffering*) OR Analges* OR Nociception*)) OR lbp OR dorsalgia OR backach* OR Lumbago*)OR SU (backpain* OR ((lumbar OR back OR spine OR sacral OR spinal) N3 (pain* OR ache* OR aching OR (Physical N1 Suffering*) OR Analges* OR Nociception*)) OR lbp OR dorsalgia OR backach* OR Lumbago*)

S2	(MH "Pain+" OR MH "Pain Measurement") AND (MH "Spine+" OR MH "Back")

S3	S1 OR S2

S4	TI (((Motor OR Movement OR kinetic OR kinematic) N1 Variabilit*) OR ((Nonlinear OR "Non linear" OR linear) N1 Dynamic*) OR ((Nonlinear OR "Non linear") N1 model) OR ((Trunk OR Postur*) N1 (control OR sway)) OR (Trunk N1 muscle* N1 activ*) OR (Movement N1 (irregularity OR alteration* OR Variabilit*)) OR (Coordinative N1 Variabilit*) OR ("Inter-segmental" N1 coordination) OR (Trunk N1 Pelvis N1 coordination*) OR (Neuromuscular N1 adaptation*) OR ((Dynamic* OR temporal) N1 structure*) OR (Temporal N1 Spatial N1 parameter*) OR (spine N1 rotational N1 stiffness) OR ((Trunk OR Dynamic) N1 (stability* OR movement OR balance))) OR AB (((Motor OR Movement OR kinetic OR kinematic) N1 Variabilit*) OR ((Nonlinear OR "Non linear" OR linear) N1 Dynamic*) OR ((Nonlinear OR "Non linear") N1 model) OR ((Trunk OR Postur*) N1 (control OR sway)) OR (Trunk N1 muscle* N1 activ*) OR (Movement N1 (irregularity OR alteration* OR Variabilit*)) OR (Coordinative N1 Variabilit*) OR ("Inter-segmental" N1 coordination) OR (Trunk N1 Pelvis N1 coordination*) OR (Neuromuscular N1 adaptation*) OR ((Dynamic* OR temporal) N1 structure*) OR (Temporal N1 Spatial N1 parameter*) OR (spine N1 rotational N1 stiffness) OR ((Trunk OR Dynamic) N1 (stability* OR movement OR balance))) OR SU (((Motor OR Movement OR kinetic OR kinematic) N1 Variabilit*) OR ((Nonlinear OR "Non linear" OR linear) N1 Dynamic*) OR ((Nonlinear OR "Non linear") N1 model) OR ((Trunk OR Postur*) N1 (control OR sway)) OR (Trunk N1 muscle* N1 activ*) OR (Movement N1 (irregularity OR alteration* OR Variabilit*)) OR (Coordinative N1 Variabilit*) OR ("Inter-segmental" N1 coordination) OR (Trunk N1 Pelvis N1 coordination*) OR (Neuromuscular N1 adaptation*) OR ((Dynamic* OR temporal) N1 structure*) OR (Temporal N1 Spatial N1 parameter*) OR (spine N1 rotational N1 stiffness) OR ((Trunk OR Dynamic) N1 (stability* OR movement OR balance)))

S5	TI (Entrop* OR (Largest N1 Lyapunov N1 Exponent*) OR (detrended N1 fluctuati* N1 analysis) OR (correlate* N1 dimension*) OR (Mutual N1 information) OR (Hurst N1 exponent*) OR (Recurrence N1 quantification N1 analysis) OR (Floquet N1 multiplier*)) OR AB (Entrop* OR (Largest N1 Lyapunov N1 Exponent*) OR (detrended N1 fluctuati* N1 analysis) OR (correlate* N1 dimension*) OR (Mutual N1 information) OR (Hurst N1 exponent*) OR (Recurrence N1 quantification N1 analysis) OR (Floquet N1 multiplier*)) OR SU (Entrop* OR (Largest N1 Lyapunov N1 Exponent*) OR (detrended N1 fluctuati* N1 analysis) OR (correlate* N1 dimension*) OR (Mutual N1 information) OR (Hurst N1 exponent*) OR (Recurrence N1 quantification N1 analysis) OR (Floquet N1 multiplier*))

S6	MH "Electromyography" **OR TI ((Center of Pressure*) OR Vicon OR OptiTrack OR (**Optotrack N1 Certus) OR (Qualisys N1 motion N1 analysis N1 system) OR (Liberty N1 Polhemus) OR (Electromagnetic N1 motion N1 track*) OR (Inertial N1 Measurement N1 Unit*) OR (motion N1 captur*)) **OR AB ((Center of Pressure*) OR Vicon OR OptiTrack OR (**Optotrack N1 Certus) OR (Qualisys N1 motion N1 analysis N1 system) OR (Liberty N1 Polhemus) OR (Electromagnetic N1 motion N1 track*) OR (Inertial N1 Measurement N1 Unit*) OR (motion N1 captur*)) **OR SU ((Center of Pressure*) OR Vicon OR OptiTrack OR (**Optotrack N1 Certus) OR (Qualisys N1 motion N1 analysis N1 system) OR (Liberty N1 Polhemus) OR (Electromagnetic N1 motion N1 track*) OR (Inertial N1 Measurement N1 Unit*) OR (motion N1 captur*))

**S7	TI (**variabilit* OR (standard N1 deviation*) OR Range OR excursion OR Mean OR Angles OR (End N1 point N1 variability*) OR (Angular N1 displac*) OR Acceleration* OR Velocit* OR (Force N1 accuracy*) OR (Total N1 displacement)) OR **AB (**variabilit* OR (standard N1 deviation*) OR Range OR excursion OR Mean OR Angles OR (End N1 point N1 variability*) OR (Angular N1 displac*) OR Acceleration* OR Velocit* OR (Force N1 accuracy*) OR (Total N1 displacement)) OR SU **(**variabilit* OR (standard N1 deviation*) OR Range OR excursion OR Mean OR Angles OR (End N1 point N1 variability*) OR (Angular N1 displac*) OR Acceleration* OR Velocit* OR (Force N1 accuracy*) OR (Total N1 displacement))

**S8	S6 AND S7**

**S9	S4 OR S5 OR S8**

**S10	S3 AND S9**

**Cochrane**

#1	(backpain* OR ((lumbar OR back OR spine OR sacral OR spinal) NEAR/3 (pain* OR ache* OR aching OR (Physical NEAR/1 Suffering*) OR Analges* OR Nociception*)) OR lbp OR dorsalgia OR backach* OR Lumbago*):ti,ab,kw

#2	(((Motor OR Movement OR kinetic OR kinematic) NEAR/1 Variabilit*) OR ((Nonlinear OR "Non linear" OR linear) NEAR/1 Dynamic*) OR ((Nonlinear OR "Non linear") NEAR/1 model) OR ((Trunk OR Postur*) NEAR/1 (control OR sway)) OR (Trunk NEAR/1 muscle* NEAR/1 activ*) OR (Movement NEAR/1 (irregularity OR alteration* OR Variabilit*)) OR (Coordinative NEAR/1 Variabilit*) OR ("Inter-segmental" NEAR/1 coordination) OR (Trunk NEAR/1 Pelvis NEAR/1 coordination*) OR (Neuromuscular NEAR/1 adaptation*) OR ((Dynamic* OR temporal) NEAR/1 structure*) OR (Temporal NEAR/1 Spatial NEAR/1 parameter*) OR (spine NEAR/1 rotational NEAR/1 stiffness) OR ((Trunk OR Dynamic) NEAR/1 (stability* OR movement OR balance))):ti,ab,kw

#3	(Entrop* OR (Largest NEAR/1 Lyapunov NEAR/1 Exponent*) OR (detrended NEAR/1 fluctuati* NEAR/1 analysis) OR (correlate* NEAR/1 dimension*) OR (Mutual NEAR/1 information) OR (Hurst NEAR/1 exponent*) OR (Recurrence NEAR/1 quantification NEAR/1 analysis) OR (Floquet NEAR/1 multiplier*)):ti,ab,kw

#4	**((Center of Pressure*) OR Vicon OR OptiTrack OR (**Optotrack NEAR/1 Certus) OR (Qualisys NEAR/1 motion NEAR/1 analysis NEAR/1 system) OR (Liberty NEAR/1 Polhemus) OR (Electromagnetic NEAR/1 motion NEAR/1 track*) OR (Inertial NEAR/1 Measurement NEAR/1 Unit*) OR (motion NEAR/1 captur*)):ti,ab,kw

**#5	(**variabilit* OR (standard NEAR/1 deviation*) OR Range OR excursion OR Mean OR Angles OR (End NEAR/1 point NEAR/1 variability*) OR (Angular NEAR/1 displac*) OR Acceleration* OR Velocit* OR (Force NEAR/1 accuracy*) OR (Total NEAR/1 displacement)):ti,ab,kw

**#6	#4 AND #5**

**S7	#2 OR #3 OR #6**

**S8	#1 AND #7**

**WOS**

#1	TS=(backpain* OR ((lumbar OR back OR spine OR sacral OR spinal) NEAR/3 (pain* OR ache* OR aching OR (Physical NEAR/1 Suffering*) OR Analges* OR Nociception*)) OR lbp OR dorsalgia OR backach* OR Lumbago*)

#2	TS=(((Motor OR Movement OR kinetic OR kinematic) NEAR/1 Variabilit*) OR ((Nonlinear OR "Non linear" OR linear) NEAR/1 Dynamic*) OR ((Nonlinear OR "Non linear")NEAR/1 model) OR ((Trunk OR Postur*) NEAR/1 (control OR sway)) OR (Trunk NEAR/1 muscle* NEAR/1 activ*) OR (Movement NEAR/1 (irregularity OR alteration* OR Variabilit*)) OR (Coordinative NEAR/1 Variabilit*) OR ("Inter-segmental" NEAR/1 coordination) OR (Trunk NEAR/1 Pelvis NEAR/1 coordination*) OR (Neuromuscular NEAR/1 adaptation*) OR ((Dynamic* OR temporal) NEAR/1 structure*) OR (Temporal NEAR/1 Spatial NEAR/1 parameter*) OR (spine NEAR/1 rotational NEAR/1 stiffness) OR ((Trunk OR Dynamic) NEAR/1 (stability* OR movement OR balance)))

#3	TS=(Entrop* OR (Largest NEAR/1 Lyapunov NEAR/1 Exponent*) OR (detrended NEAR/1 fluctuati* NEAR/1 analysis) OR (correlate* NEAR/1 dimension*) OR (Mutual NEAR/1 information) OR (Hurst NEAR/1 exponent*) OR (Recurrence NEAR/1 quantification NEAR/1 analysis) OR (Floquet NEAR/1 multiplier*))

#4	TS=**((Center of Pressure*) OR Vicon OR OptiTrack OR (**Optotrack NEAR/1 Certus) OR (Qualisys NEAR/1 motion NEAR/1 analysis NEAR/1 system) OR (Liberty NEAR/1 Polhemus) OR (Electromagnetic NEAR/1 motion NEAR/1 track*) OR (Inertial NEAR/1 Measurement NEAR/1 Unit*) OR (motion NEAR/1 captur*))

**#5	TS=(**variabilit* OR (standard NEAR/1 deviation*) OR Range OR excursion OR Mean OR Angles OR (End NEAR/1 point NEAR/1 variability*) OR (Angular NEAR/1 displac*) OR Acceleration* OR Velocit* OR (Force NEAR/1 accuracy*) OR (Total NEAR/1 displacement))

**#6	#4 AND #5**

**S7	#2 OR #3 OR #6**

**S8	#1 AND #7**

**SportDiscus**

S1	DE "BACKACHE" OR DE "LUMBAR pain" OR DE "BACKACHE exercise therapy" OR TI (backpain* OR ((lumbar OR back OR spine OR sacral OR spinal) N3 (pain* OR ache* OR aching OR (Physical N1 Suffering*) OR Analges* OR Nociception*)) OR lbp OR dorsalgia OR backach* OR Lumbago*) OR AB (backpain* OR ((lumbar OR back OR spine OR sacral OR spinal) N3 (pain* OR ache* OR aching OR (Physical N1 Suffering*) OR Analges* OR Nociception*)) OR lbp OR dorsalgia OR backach* OR Lumbago*)OR SU (backpain* OR ((lumbar OR back OR spine OR sacral OR spinal) N3 (pain* OR ache* OR aching OR (Physical N1 Suffering*) OR Analges* OR Nociception*)) OR lbp OR dorsalgia OR backach* OR Lumbago*)

S2	(DE "PAIN" OR DE "CHRONIC pain" OR DE "PAIN management" OR DE "PAIN measurement") AND (DE "SPINE" OR DE "BACK")

S3	S1 OR S2

S4	TI (((Motor OR Movement OR kinetic OR kinematic) N1 Variabilit*) OR ((Nonlinear OR "Non linear" OR linear) N1 Dynamic*) OR ((Nonlinear OR "Non linear") N1 model) OR ((Trunk OR Postur*) N1 (control OR sway)) OR (Trunk N1 muscle* N1 activ*) OR (Movement N1 (irregularity OR alteration* OR Variabilit*)) OR (Coordinative N1 Variabilit*) OR ("Inter-segmental" N1 coordination) OR (Trunk N1 Pelvis N1 coordination*) OR (Neuromuscular N1 adaptation*) OR ((Dynamic* OR temporal) N1 structure*) OR (Temporal N1 Spatial N1 parameter*) OR (spine N1 rotational N1 stiffness) OR ((Trunk OR Dynamic) N1 (stability* OR movement OR balance))) OR AB (((Motor OR Movement OR kinetic OR kinematic) N1 Variabilit*) OR ((Nonlinear OR "Non linear" OR linear) N1 Dynamic*) OR ((Nonlinear OR "Non linear") N1 model) OR ((Trunk OR Postur*) N1 (control OR sway)) OR (Trunk N1 muscle* N1 activ*) OR (Movement N1 (irregularity OR alteration* OR Variabilit*)) OR (Coordinative N1 Variabilit*) OR ("Inter-segmental" N1 coordination) OR (Trunk N1 Pelvis N1 coordination*) OR (Neuromuscular N1 adaptation*) OR ((Dynamic* OR temporal) N1 structure*) OR (Temporal N1 Spatial N1 parameter*) OR (spine N1 rotational N1 stiffness) OR ((Trunk OR Dynamic) N1 (stability* OR movement OR balance))) OR SU (((Motor OR Movement OR kinetic OR kinematic) N1 Variabilit*) OR ((Nonlinear OR "Non linear" OR linear) N1 Dynamic*) OR ((Nonlinear OR "Non linear") N1 model) OR ((Trunk OR Postur*) N1 (control OR sway)) OR (Trunk N1 muscle* N1 activ*) OR (Movement N1 (irregularity OR alteration* OR Variabilit*)) OR (Coordinative N1 Variabilit*) OR ("Inter-segmental" N1 coordination) OR (Trunk N1 Pelvis N1 coordination*) OR (Neuromuscular N1 adaptation*) OR ((Dynamic* OR temporal) N1 structure*) OR (Temporal N1 Spatial N1 parameter*) OR (spine N1 rotational N1 stiffness) OR ((Trunk OR Dynamic) N1 (stability* OR movement OR balance)))

S5	TI (Entrop* OR (Largest N1 Lyapunov N1 Exponent*) OR (detrended N1 fluctuati* N1 analysis) OR (correlate* N1 dimension*) OR (Mutual N1 information) OR (Hurst N1 exponent*) OR (Recurrence N1 quantification N1 analysis) OR (Floquet N1 multiplier*)) OR AB (Entrop* OR (Largest N1 Lyapunov N1 Exponent*) OR (detrended N1 fluctuati* N1 analysis) OR (correlate* N1 dimension*) OR (Mutual N1 information) OR (Hurst N1 exponent*) OR (Recurrence N1 quantification N1 analysis) OR (Floquet N1 multiplier*)) OR SU (Entrop* OR (Largest N1 Lyapunov N1 Exponent*) OR (detrended N1 fluctuati* N1 analysis) OR (correlate* N1 dimension*) OR (Mutual N1 information) OR (Hurst N1 exponent*) OR (Recurrence N1 quantification N1 analysis) OR (Floquet N1 multiplier*))

S6	DE "ELECTROMYOGRAPHY" **OR TI ((Center of Pressure*) OR Vicon OR OptiTrack OR (**Optotrack N1 Certus) OR (Qualisys N1 motion N1 analysis N1 system) OR (Liberty N1 Polhemus) OR (Electromagnetic N1 motion N1 track*) OR (Inertial N1 Measurement N1 Unit*) OR (motion N1 captur*)) **OR AB ((Center of Pressure*) OR Vicon OR OptiTrack OR (**Optotrack N1 Certus) OR (Qualisys N1 motion N1 analysis N1 system) OR (Liberty N1 Polhemus) OR (Electromagnetic N1 motion N1 track*) OR (Inertial N1 Measurement N1 Unit*) OR (motion N1 captur*)) **OR SU ((Center of Pressure*) OR Vicon OR OptiTrack OR (**Optotrack N1 Certus) OR (Qualisys N1 motion N1 analysis N1 system) OR (Liberty N1 Polhemus) OR (Electromagnetic N1 motion N1 track*) OR (Inertial N1 Measurement N1 Unit*) OR (motion N1 captur*))

**S7	TI (**variabilit* OR (standard N1 deviation*) OR Range OR excursion OR Mean OR Angles OR (End N1 point N1 variability*) OR (Angular N1 displac*) OR Acceleration* OR Velocit* OR (Force N1 accuracy*) OR (Total N1 displacement)) OR **AB (**variabilit* OR (standard N1 deviation*) OR Range OR excursion OR Mean OR Angles OR (End N1 point N1 variability*) OR (Angular N1 displac*) OR Acceleration* OR Velocit* OR (Force N1 accuracy*) OR (Total N1 displacement)) OR SU **(**variabilit* OR (standard N1 deviation*) OR Range OR excursion OR Mean OR Angles OR (End N1 point N1 variability*) OR (Angular N1 displac*) OR Acceleration* OR Velocit* OR (Force N1 accuracy*) OR (Total N1 displacement))

**S8	S6 AND S7**

**S9	S4 OR S5 OR S8**

**S10	S3 AND S9**

**Number of articles	23-5-2022**

Cinahl	1040

Cochrane	344

Embase	1412

Medline	1738

PubMed	1595

SportDiscus	658

WOS	1727

**Total before removal duplicates	8514**

**Total after removal duplicates	3796**
