## Supplementary material for "Not all movements are equal: Differences in variability of trunk motor behavior between people with and without low back pain - A Systematic Review": S1 File

To enable PROSPERO to focus on COVID-19 submissions, this registration record has undergone basic automated checks for eligibility and is published exactly as submitted. PROSPERO has never provided peer review, and usual checking by the PROSPERO team does not endorse content. Therefore, automatically published records should be treated as any other PROSPERO registration. Further detail is provided here.

### Citation

Florian Abu Bakar, Bart Staal, Robert van Cingel, Raymond Ostelo, Jaap van Dieën. Not all movements are equal: Differences in magnitude and structure of movement variability between people with and without low back pain - A Systematic Review. PROSPERO 2020 CRD42020180003 Available from:
https://www.crd.york.ac.uk/prospero/display_record.php?ID=CRD42020180003

### Review question

Are there differences in movement variability regarding size and structure between people with and without low back pain?

### Searches

Sources:

CINAHL

PubMed

Embase

Cochrane

Web of Science

SPORTDiscus

### Types of study to be included

Eligibility criteria of type of studies:

- We will include movement lab-based studies, case-control studies and Randomized Control Trials (baseline measurements only) that measure the concept of movement variability of the spine

- Studies must include both, a LBP group and a control group; Studies without control group are excluded

- Reviews of any kind are excluded

- There is no exclusion criteria regarding the number of participants in each study

### Condition or domain being studied

Low-back pain is a widely prevalent condition causing a high burden of disease globally and is associated with high economic costs. Unfortunately, effective treatment options are scarce. Movement variability, a hallmark of human movement, describes the phenomenon that no single activity is performed in exactly the same manner. Literature suggests movement variability might be altered in patients with low-back pain, presumably due to perceived or actual risks of pain provocation and loss of movement control.

### Participants/population

Eligibility criteria of participants:

- Inclusion: Adults (above 17 years of age), acute low-back pain, subacute low-back. pain, chronic low-back pain, non-specific low-back pain

- Exclusion: Adolescents (under 18 years of age), specific low-back (e.g. xxx), and post-surgery.

### Intervention(s), exposure(s)

not applicable - There is neither an intervention nor exposures

### Comparator(s)/control

Main outcome: Differences of movement variability

Central Determinant: participants with and without low-back pain

### Main outcome(s)

Main outcome: Differences of movement variability

#### Measures of effect

global method to measure movement variability (magnitude or structure)

specific measure for variability (e.g. standard deviation, sample entropy etc.)

### Additional outcome(s)

none

#### Measures of effect

not applicable

### Data extraction (selection and coding)

Study selection:

The reviewers will independently screen the abstracts and titles retrieved by the search strategy applying the criteria for eligibility. Duplicate articles are excluded. Full texts will be obtained when the abstract fulfills the inclusion criteria and will subsequently be screened for eligibility. Studies will be independently classified as included, excluded or undecided using Rayyan. Where no abstract is available, full-text articles will be obtained unless the article can be confidently excluded by its title alone. In general, if there is any doubt about exclusion of the study, the study will proceed to full-text screening to reduce the likelihood of an incorrect exclusion. Disagreements will be solved by discussion and consensus between the two reviewers. Where no consensus can be reached, the third reviewer will arbitrate.

Data extraction:

Data will be independently extracted from full texts by two review authors. Any disagreements will be discussed between the two authors and a third review author will be consulted if necessary.

The following data are extracted:

1. General characteristics of the studies (e.g. Author, year of publication, title and study design...)

2. Characteristics of the participants (e.g. Number of participants, age, gender, study population details...)

3. Description of: movement tasks, used aperture/instruments, global method to measure movement variability (magnitude or structure), specific measure for variability (e.g. standard deviation, sample entropy) and the authors' reported results and conclusions

### Risk of bias (quality) assessment

Risk of Bias will be assessed by two 2 independent reviewers. When necessary, a third reviewer will be contacted. Risk of bias will be assessed with the Quality in Prognosis Studies (QUIPS) tool. The QUIPS tool considers the following 6 domains of bias:

Bias due to

1.Study participation

2.Study attrition

3.Prognostic factor measurement

4.Outcome measurement

5.Study confounding and

6.Statistical analysis and reporting.

Due to the nature of this review, risk of bias will be assessed in 5 of the 6 domains proposed in the QUIPS tool. The domain 'study attrition' is not applicable due to the cross-sectional character of the studies to be included. Within each domain, the three to seven issues are scored. Possible responses are: `yes', `partial', `no' or `unsure'.

The responses on these items are combined to assess the risk of bias per domain. The risk of bias for each domain will be classified as `high' (+), `moderate' (+/−) or `low' (−) risk of bias. The overall risk of bias will not be reported.

### Strategy for data synthesis

A narrative synthesis of the included studies will be provided including tables of study characteristics, participants, intervention details, and relevant outcomes. Appropriate statistical techniques will be used. If applicable, this review will also include a meta-analysis to calculate pooled effect sizes across studies.

### Analysis of subgroups or subsets

If the literature included allows it, we will perform subgroup analyses on factors such as:

- duration and severity of low back pain of subjects and its influence on the variability measure of interest

- age subjects and its influence on the variability measure of interest

- psychosocial variables (such as fear of movement or catastrophizing) of subjects and its influence on the variability measure of interest

- place of recruitment of subjects and its influence on the variability measure of interest (e.g. outpatients, local practices, etc.)

### Contact details for further information

Florian Abu Bakar

### Organisational affiliation of the review

HAN University of Applied Sciences

https://www.han.nl/

### Review team members and their organisational affiliations

Mr Florian Abu Bakar. HAN University of Applied Sciences

Dr Bart Staal. HAN University of Applied Sciences

Dr Robert van Cingel. Sport Medisch Centrum Papendal

Professor Raymond Ostelo. Vrije Universiteit Amsterdam

Professor Jaap van Dieën. Vrije Universiteit Amsterdam

### Collaborators

Hiroko Saito. Department of physical therapy, School of Health Sciences, Tokyo University of Technology, Tokyo, Japan

### Type and method of review

Methodology, Systematic review

### Anticipated or actual start date

01 September 2020

### Anticipated completion date

31 January 2021

### Funding sources/sponsors

This review is funded by the 'Nederlandse Organisatie voor Wetenschappelijk Onderzoek ' (NWO)

### Grant number(s)

State the funder, grant or award number and the date of award

Project number: 023.011.018

### Conflicts of interest

Funding disclosures: Florian Abu Bakar was funded by a doctoral grand for teachers from

the Dutch Organization for Scientific Research (NWO). The other authors did not receive financial support for this study.

Yes

### Language

English

### Country

Netherlands

### Stage of review

Review Ongoing

### Subject index terms status

Subject indexing assigned by CRD

### Subject index terms

Biomechanical Phenomena; Humans; Low Back Pain; Lumbosacral Region

### Date of registration in PROSPERO

20 September 2020

### Date of first submission

20 August 2020

### Stage of review at time of this submission

| Stage | Started | Completed |
|---|---|---|
| Preliminary searches | Yes | No |
| Piloting of the study selection process | No | No |
| Formal screening of search results against eligibility criteria | No | No |
| Data extraction | No | No |
| Risk of bias (quality) assessment | No | No |
| Data analysis | No | No |

*The record owner confirms that the information they have supplied for this submission is accurate and complete and they understand that deliberate provision of inaccurate information or omission of data may be construed as scientific misconduct.*

*The record owner confirms that they will update the status of the review when it is completed and will add publication details in due course.*

### Versions

20 September 2020
